# Inflammatory sub-phenotypes in sepsis: relationship to outcomes, treatment effect and transcriptomic sub-phenotypes

**DOI:** 10.1101/2022.07.12.22277463

**Authors:** David B Antcliffe, Yuxin Mi, Shalini Santhakumaran, Katie L Burnham, A Toby Prevost, Josie K Ward, Timothy Marshall, Claire Bradley, Farah Al-Beidh, Paula Hutton, Stuart McKechnie, Emma E Davenport, Charles J Hinds, Cecilia M O’Kane, Daniel F McAuley, Manu Shankar-Hari, Anthony C Gordon, Julian C Knight

## Abstract

**Rationale:** Heterogeneity of sepsis limits discovery and targeting of treatments. Clustering approaches in critical illness have identified patient groups who may respond differently to therapies. These include in acute respiratory distress syndrome (ARDS) two inflammatory sub-phenotypes, using latent class analysis (LCA), and in sepsis two Sepsis Response Signatures (SRS), based on transcriptome profiling. It is unknown if inflammatory sub-phenotypes such as those identified in ARDS are present in sepsis and how sub-phenotypes defined with different techniques compare.

**Objectives:** To identify inflammatory sub-phenotypes in sepsis using LCA and assess if these show differential treatment responses. These sub-phenotypes were compared to hierarchical clusters based on inflammatory mediators and to SRS sub-phenotypes.

**Methods:** LCA was applied to clinical and biomarker data from two septic shock randomized trials. VANISH compared norepinephrine to vasopressin and hydrocortisone to placebo and LeoPARDS compared levosimendan to placebo. Hierarchical cluster analysis (HCA) was applied to 65, 21 and 11 inflammatory mediators measured in patients from the GAinS (n=124), VANISH (n=155) and LeoPARDS (n=484) studies.

**Measurements and Main Results:** LCA and HCA identified a sub-phenotype of patients with high cytokine levels and worse organ dysfunction and survival, with no interaction between LCA classes and trial treatment responses. Comparison of inflammatory and transcriptomic sub-phenotypes revealed some similarities but without sufficient overlap that they are interchangeable.

**Conclusions:** A sub-phenotype with high levels of inflammation and increased disease severity is consistently identifiable in sepsis, with similarities to that described in ARDS. There was limited overlap with the transcriptomic sub-phenotypes.

## Introduction

Sepsis, life threatening organ dysfunction caused by a dysregulated host response to infection (1), causes 11 million deaths worldwide, annually (2). Sepsis is a heterogeneous syndrome, which is a barrier to finding novel therapies (3–5), prompting a search for sepsis sub-populations (also referred to as sub-phenotypes), with the hope that patients could be stratified for targeted treatments.

Sub-phenotypes have been reported in sepsis (6–8) and ARDS (9–13). In ARDS latent class analysis (LCA) using clinical and biomarker data, has consistently identified two sub-phenotypes. A hyper-inflammatory sub-phenotype is characterized by higher cytokine levels, shock, worse acidosis, worse clinical outcomes and potentially different responses to treatment compared to the hypo-inflammatory sub-phenotype (9–13). The sub-phenotypes in ARDS are predominantly driven by differences in inflammatory mediators, yet in sepsis there has not yet been an attempt to apply LCA in this way (14).

We have previously reported two sepsis sub-phenotypes from genome-wide gene expression profiling of peripheral blood leukocytes (Sepsis Response Signatures – SRS) in patients admitted to intensive care units (ICU) with community acquired pneumonia (20) or fecal peritonitis (21). Based on differential gene expression, SRS1 shows features of immune suppression compared with SRS2 and is associated with higher mortality. SRS sub-phenotypes have shown potential to identify patients with septic shock who respond differently to hydrocortisone (22) with this intervention increasing mortality in SRS2 patients but not in SRS1. It is unknown how these transcriptomic sub-phenotypes relate to other sub-populations based on clustering of alternative data types.

To fully appreciate the utility of sub-phenotypes it is important to understand if they are specific to particular syndromes, such as ARDS, or are present in all critically ill patients with sepsis. It is also important to understand the relationship between sub-phenotypes defined using different types of data so that the optimal method can be used to inform a precision medicine approach to treatment.

We hypothesized that LCA of clinical and inflammatory biomarker data would identify distinct sepsis sub-phenotypes which might respond differently to levosimendan, hydrocortisone, or vasopressin and that LCA defined sub-phenotypes would be distinct from those defined by clustering of only inflammatory or transcriptomic data. Some of these results have been reported as an abstract (23).

## Methods

Detailed methodology can be found in the supplement.

### Patients

Blood samples and clinical data were available from the VANISH (24) and LeoPARDS (25) septic shock trials, which included patients with sepsis from any source, and the Genomic Advances in Sepsis (GAinS) (20, 26) study, which included patients with sepsis due to community acquired pneumonia or fecal peritonitis. All had ethics approval and informed written consent was obtained from patients or their legal representatives.

VANISH was a factorial (2×2) randomized trial comparing vasopressin to norepinephrine and hydrocortisone to placebo in septic shock. Biomarker assessment was available for 176/409 patients. The LeoPARDS trial was a randomized trial comparing levosimendan to placebo in septic shock. Biomarker assessment was available for 493/516 patients. GAinS was an observational study of patients admitted to ICU with sepsis or septic shock. Biomarker assessment was performed on 124 patients.

### Biomarker Measurement

Plasma was collected on the first day of ICU admission (GAinS) or of septic shock (VANISH and LeoPARDS). Sixty-five inflammatory mediators were measured in GAinS, twenty-one in VANISH and eleven in LeoPARDS (Table S1). Biomarkers including the inflammatory mediators and other blood marker molecules were measured as described in the supplement.

### Transcriptomic Sub-phenotypes

Genome-wide transcriptomic data in blood leukocytes, acquired using Illumina Human-HT-12 v4 Expression BeadChips, were available from the VANISH and GAinS studies. SRS sub-phenotypes have previously been assigned in the GAinS patients using unsupervised clustering on genome-wide transcriptomic data or a predictive 7-gene model, and in VANISH patients using the 7-gene model (20–22).

### Latent Class Analysis

LCA was performed separately on the data from the two clinical trials, LeoPARDS and VANISH, agnostic of outcome. These datasets were used for LCA as both had the same biomarkers and clinical variables available and, being interventional trials, allowed exploration of sub-phenotype and treatment interaction. Variables were chosen for inclusion based on their associations with sepsis pathophysiology (PaO_2_/FiO_2_ ratio, creatinine, platelets, bilirubin, lactate, IL-1β, IL-6, IL-8, IL-10, IL-17, IL-18, MPO, sICAM, ANG-2, sTNFr, MCP-1 (CCL2), troponin and NT-proBNP). The acute physiology element of the APACHE II (APS-APII) score was included as a covariate as we expected this severity score to be associated with the biomarkers independently of subclass. Other baseline clinical and demographic variables which may be predictive of subclass were included in the models as class predictors. Biomarkers were log transformed and standardized due to skewness.

Observations outside the limits of detection were treated as censored (27). Models were fit using the gsem package in Stata 15 (StataCorp. 2017. Stata Statistical Software: Release 15. College Station, TX: StataCorp LLC).

### Clinical Outcomes

Differential treatment response based on LCA-defined sub-phenotypes were explored using the following outcomes. The primary outcome for the LeoPARDS trial data in this analysis, was survival at 3 months. Mean total SOFA score over 28 days (or ICU stay, whichever was shorter) and 28-day survival were examined as secondary outcomes. For the VANISH trial, we examined 28-day survival, as 3-month survival was not available. Survival free of renal failure to 28 days amongst patients not in renal failure at baseline, and days alive and free of renal failure up to 28 days for all other patients were also examined.

### Hierarchical Cluster Analysis

To understand how LCA defined clusters compared to those derived using a different clustering approach using only inflammatory mediator data, hierarchical cluster analysis (HCA) was applied to the inflammatory mediator panels in VANISH and LeoPARDs independently. As HCA is unable to handle missing data, only patients with complete panels were included. To facilitate comparison of inflammatory sub-phenotypes to previously defined SRS groups, HCA was also applied to the panel of inflammatory mediators measured in GAinS. Clustering was performed using the hclust function in R (28). The optimal number of clusters was defined by inspection of the dendrograms, comparison of test sample distances in cross-validation, and by determining cluster robustness by consensus clustering (29).

### Differential Gene Expression

Microarray data from GAinS and VANISH were co-normalized using the vsn package (30) and batch corrected using the ComBat function from the sva package (31) in R, resulting in 28220 communal probes after quality control. Following additional quality control with MixupMapper (32), baseline gene expression data were available for 115 patients in GAinS and 149 in VANISH from those included in the inflammatory mediator HCA. Genes differentially expressed between sub-phenotypes were identified using the limma package (33).

### Statistical Analysis

Data were compared using the Mann-Whitney U, Kruskal-Wallis, Chi-squared or Fisher’s exact test (Fisher’s exact test was used when the number of events was <10) as appropriate. The Benjamini-Hochberg procedure was applied to comparisons of inflammatory mediators between HCA and SRS groups, but as this was an exploratory analysis p-values for clinical comparisons were not corrected for multiple comparisons. All tests were two-sided and a p-value or FDR <0.05 was taken as statistical significance. Statistical analysis was performed in R (28) and SPSS version 25 (IBM, USA).

## Results

### Latent Class Analysis

For LeoPARDS, entropy was over 0.85 for all models and mean class probabilities were over 0.90, indicating good class separation. The AIC and BIC decreased as the number of classes increased without reaching a minimum value, but the improvement from 3 to 4 classes was much smaller and models with more than four classes did not converge (Table S2, Table S3, Figure S1) so the simpler 3-class model was selected. The variables showing the greatest separation between classes were IL-6, IL-8, MCP-1(CCL2), IL-10 and IL-1β (Figure 1A).

**Figure 1.**
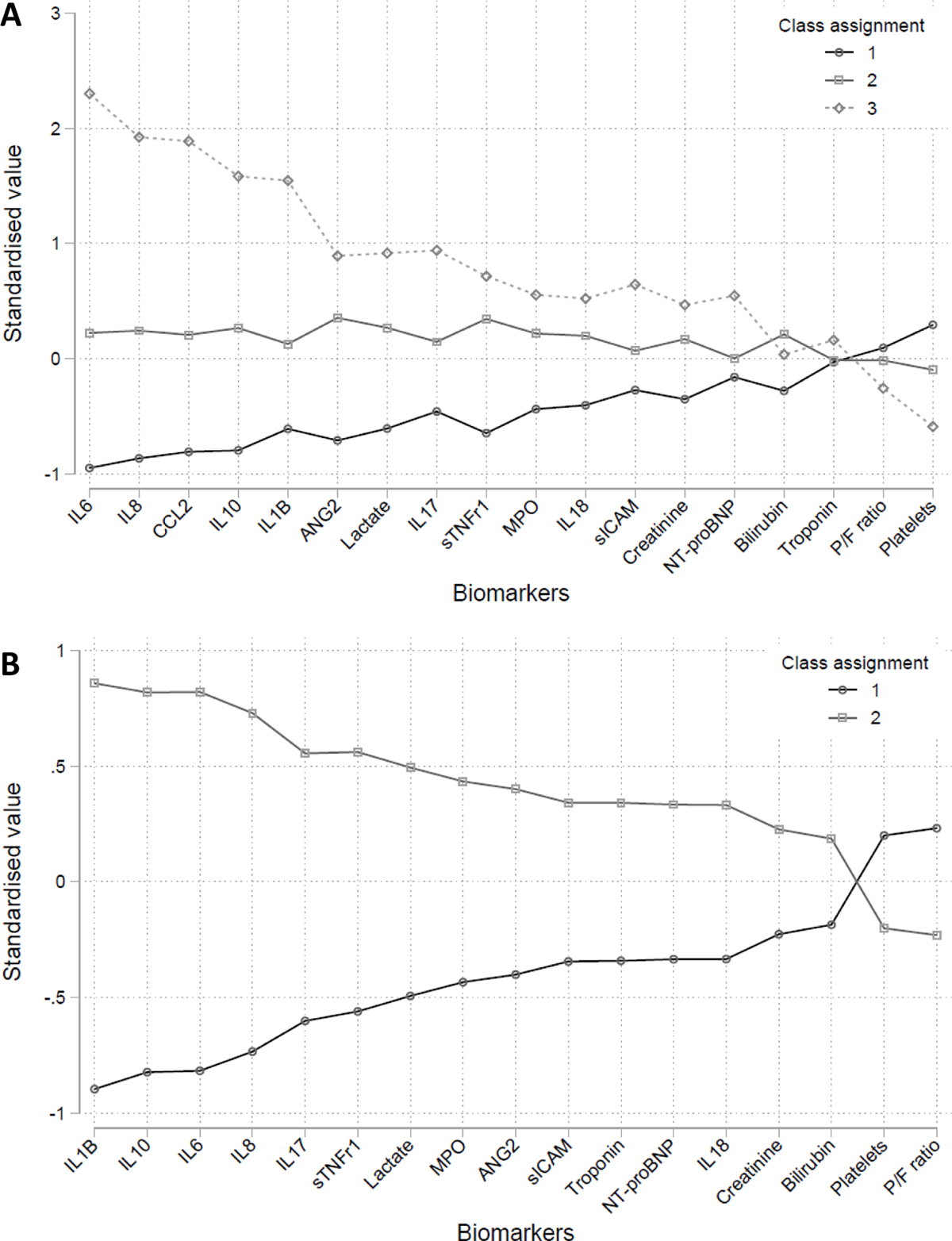
Estimated latent class analysis (LCA) class means of each indicator by class, A) LeoPARDS trial B) VANISH trial. Indicators have been standardized to ensure zero population mean and facilitate comparison. Indicators are ordered by the difference between classes for almost all indicators. In the LeoPARDS trial indicators are ordered by the difference between class 1 and class 3, the estimated means for class 2 were in between those for classes 1 and 3.

Around half the patients were assigned to class 2 (n=247), with 191 to class 1 and 55 to class 3. Baseline clinical characteristics, outcomes and biomarker concentrations by class are shown in Tables 1 and 2. Sensitivity analyses gave comparable results with 94% agreement in class assignment (Table S4). Survival varied by class (p=0.001) and was lowest in class 3 (41.8% (23/55)) compared to other classes (69.8% (132/189) in class 1; 63.0% (155/246) in class 2). A multinomial logit model with IL-6, IL-8, IL-10 and MCP-1 as predictors gave a sensitivity of around 0.9, and a specificity of 0.9 or over for all classes (Figure S2, Table S5). No treatment sub-phenotype interaction was seen for any of the outcomes in the LeoPARDS trial (Figure 2).

**Figure 2.**
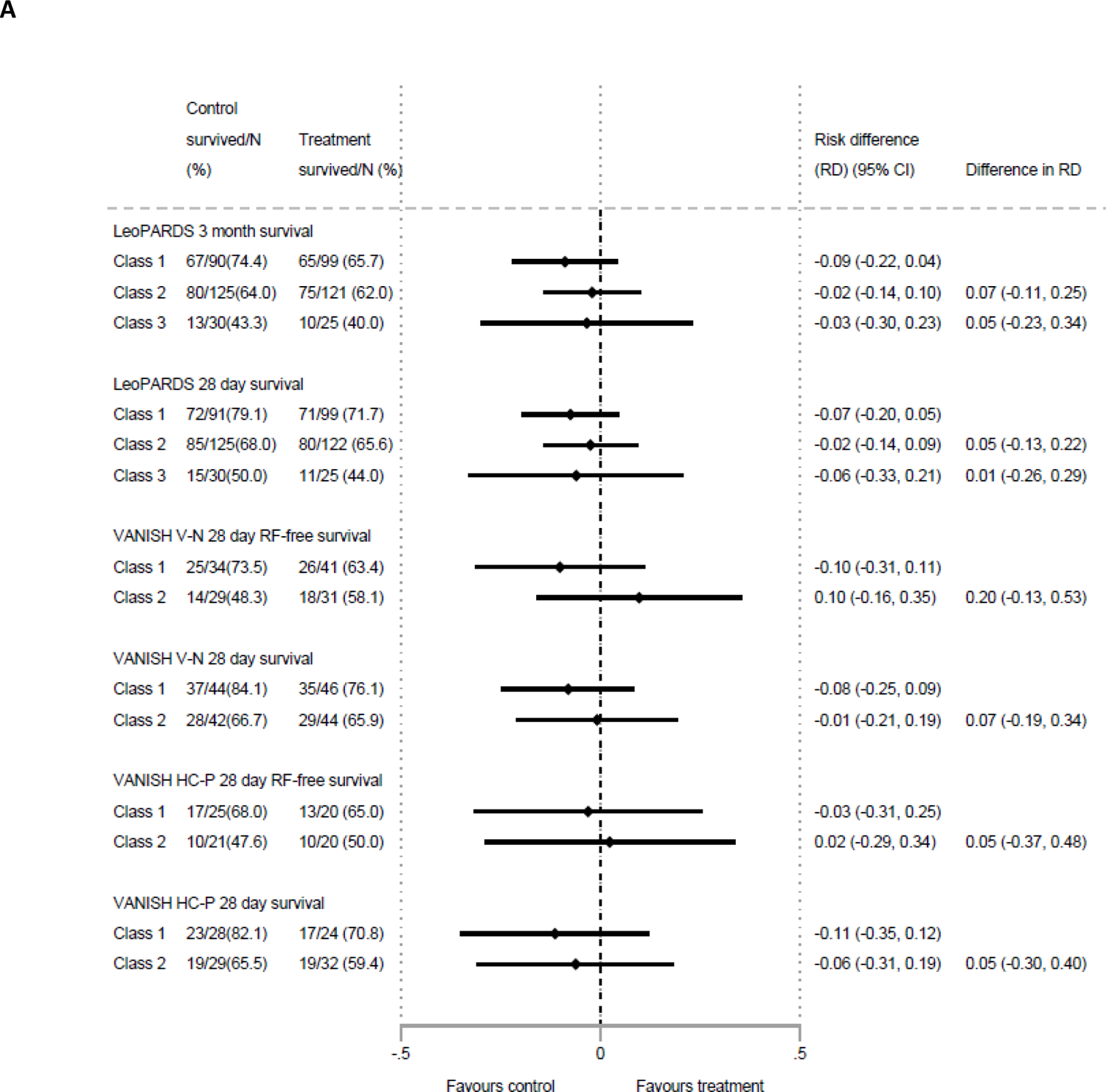
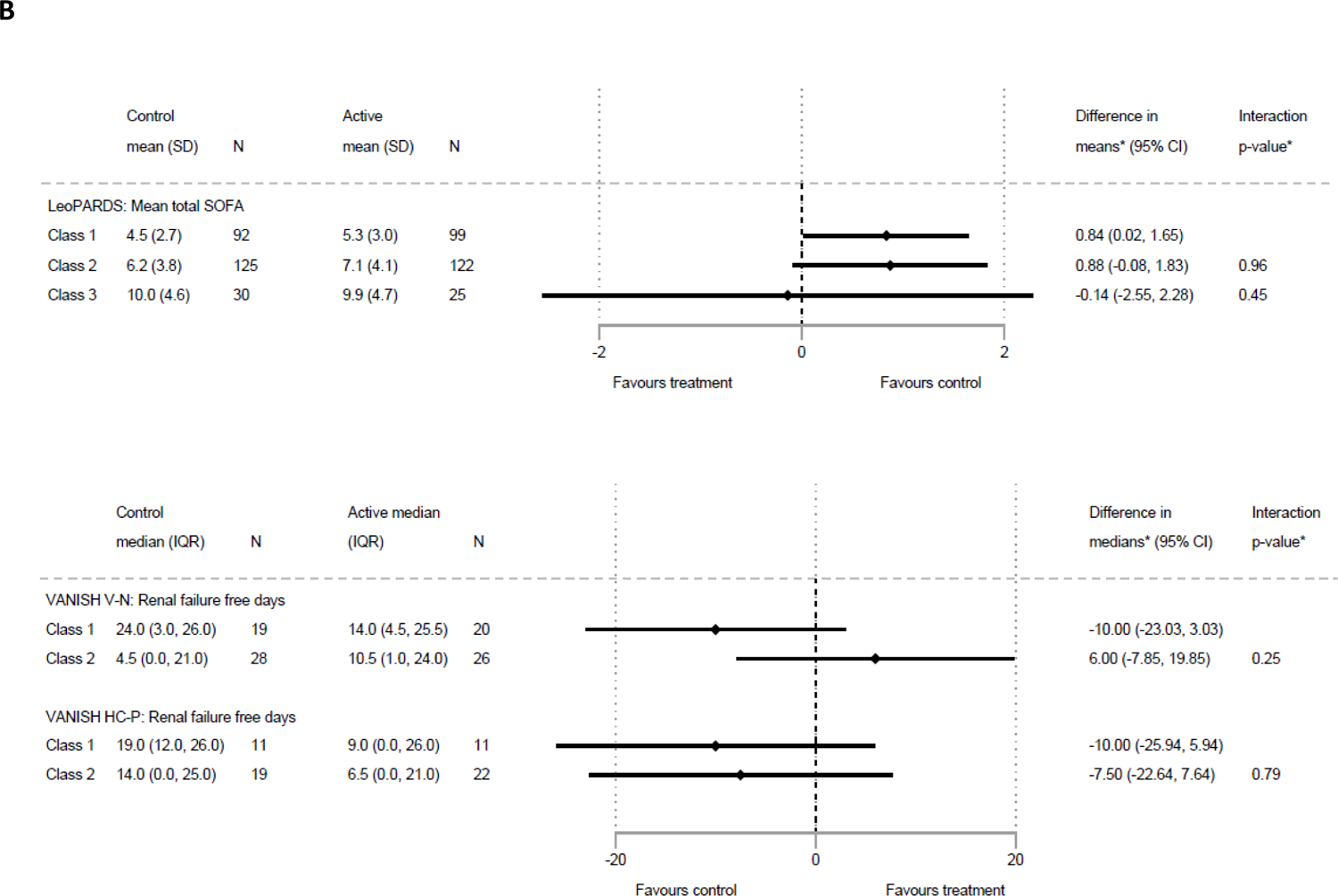
A) Forest plot showing treatment differences by latent class and trial for binary outcomes. LeoPARDS trial: 3-month survival, 28-day survival. VANISH trial: 28-day survival, 28-day survival free of renal failure. (V-N, vasopressin – norepinephrine; HC-P = hydrocortisone-placebo; RF, renal failure); B) Forest plot showing treatment differences by latent class and trial for continuous outcomes. LeoPARDS trial: mean daily SOFA score. VANISH: renal failure free days. (V-N, vasopressin – norepinephrine; HC-P = hydrocortisone-placebo)

**Table 1.**
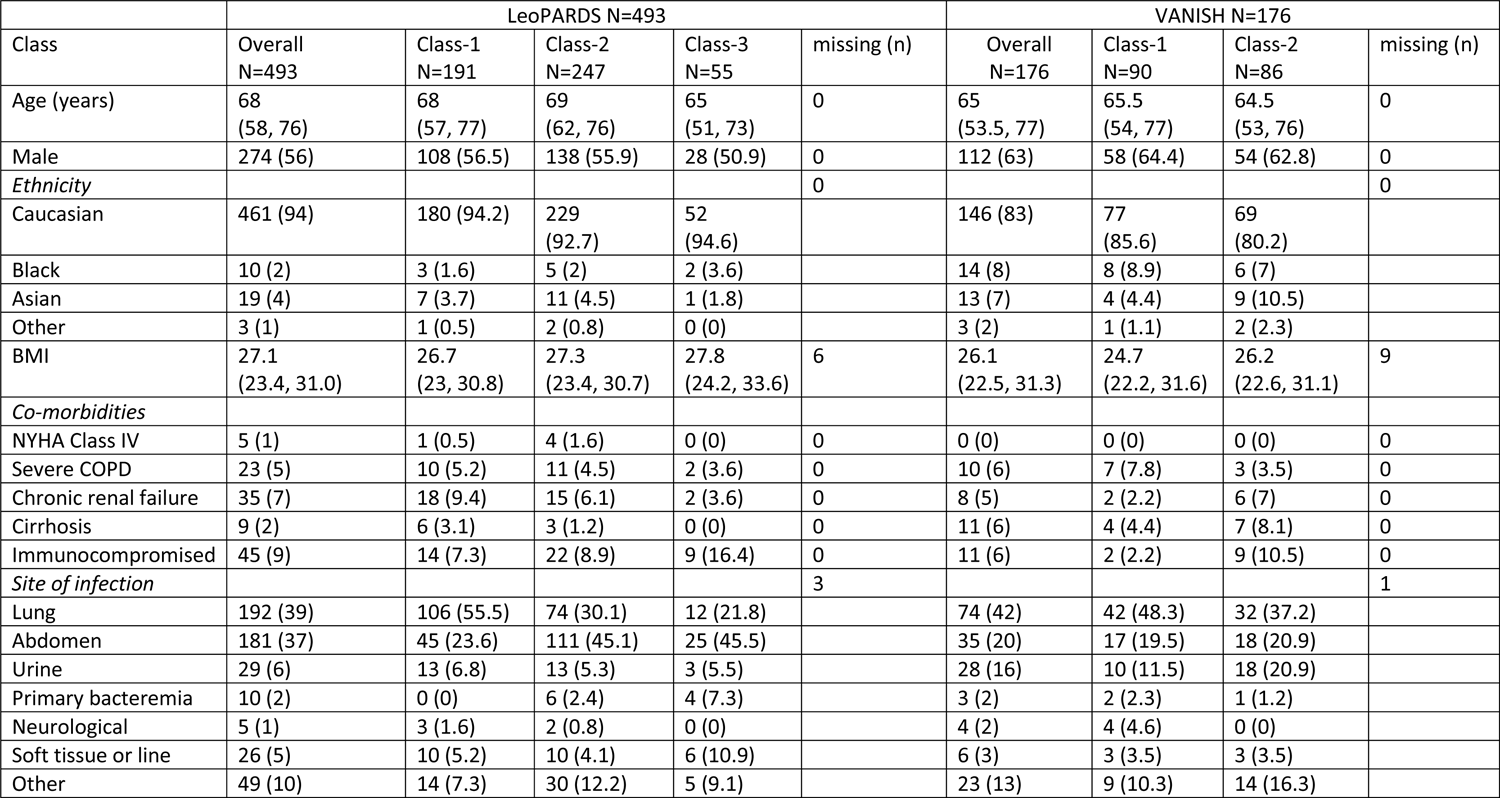
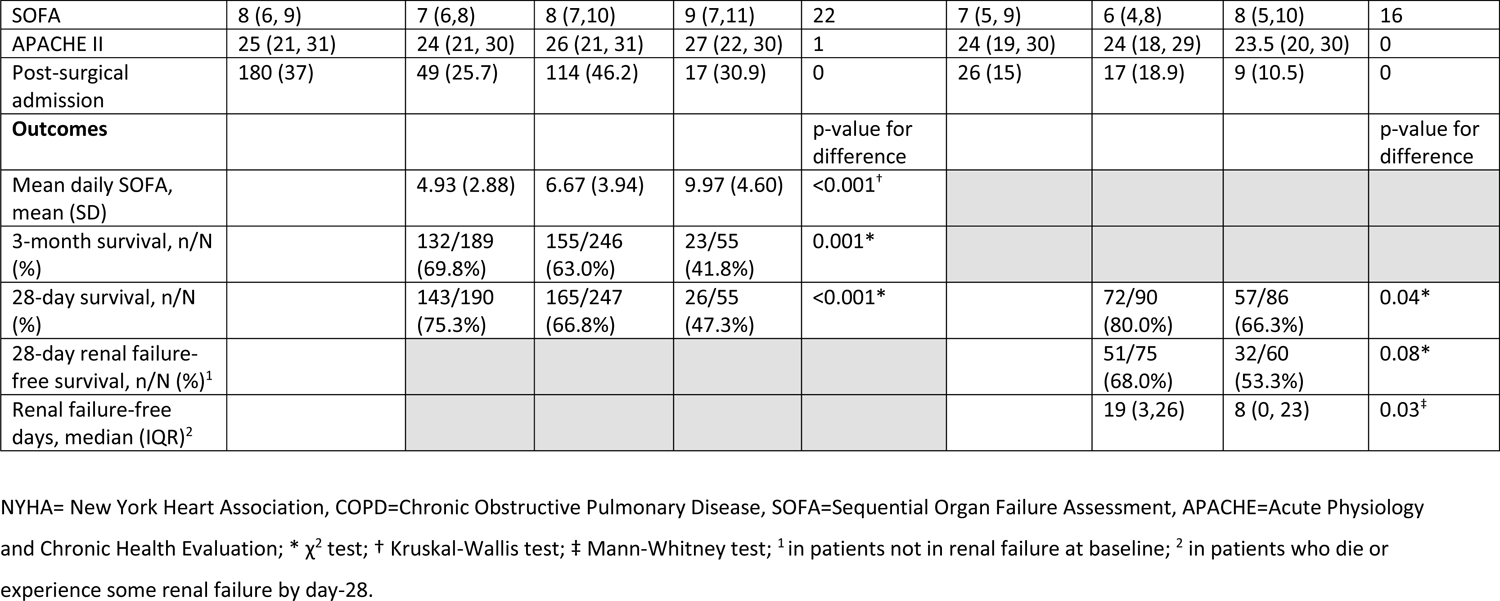
Baseline characteristics and outcomes overall and by LCA assigned class. Values are median (interquartile range) for continuous variables and n (%) for categorical variables.

|  | LeoPARDS N=493 |  |  |  |  | VANISH N=176 |  |  |  |
| --- | --- | --- | --- | --- | --- | --- | --- | --- | --- |
| Class | Overall<br>N=493 | Class-1<br>N=191 | Class-2<br>N=247 | Class-3<br>N=55 | missing (n) | Overall<br>N=176 | Class-1<br>N=90 | Class-2<br>N=86 | missing (n) |
| Age (years) | 68<br>(58, 76) | 68<br>(57, 77) | 69<br>(62, 76) | 65<br>(51, 73) | 0 | 65<br>(53.5, 77) | 65.5<br>(54, 77) | 64.5<br>(53, 76) | 0 |
| Male | 274 (56) | 108 (56.5) | 138 (55.9) | 28 (50.9) | 0 | 112 (63) | 58 (64.4) | 54 (62.8) | 0 |
| *Ethnicity* |  |  |  |  | 0 |  |  |  | 0 |
| Caucasian | 461 (94) | 180 (94.2) | 229<br>(92.7) | 52<br>(94.6) |  | 146 (83) | 77<br>(85.6) | 69<br>(80.2) |  |
| Black | 10 (2) | 3 (1.6) | 5 (2) | 2 (3.6) |  | 14 (8) | 8 (8.9) | 6 (7) |  |
| Asian | 19 (4) | 7 (3.7) | 11 (4.5) | 1 (1.8) |  | 13 (7) | 4 (4.4) | 9 (10.5) |  |
| Other | 3 (1) | 1 (0.5) | 2 (0.8) | 0 (0) |  | 3 (2) | 1 (1.1) | 2 (2.3) |  |
| BMI | 27.1<br>(23.4, 31.0) | 26.7<br>(23, 30.8) | 27.3<br>(23.4, 30.7) | 27.8<br>(24.2, 33.6) | 6 | 26.1<br>(22.5, 31.3) | 24.7<br>(22.2, 31.6) | 26.2<br>(22.6, 31.1) | 9 |
| *Co-morbidities* |  |  |  |  |  |  |  |  |  |
| NYHA Class IV | 5 (1) | 1 (0.5) | 4 (1.6) | 0 (0) | 0 | 0 (0) | 0 (0) | 0 (0) | 0 |
| Severe COPD | 23 (5) | 10 (5.2) | 11 (4.5) | 2 (3.6) | 0 | 10 (6) | 7 (7.8) | 3 (3.5) | 0 |
| Chronic renal failure | 35 (7) | 18 (9.4) | 15 (6.1) | 2 (3.6) | 0 | 8 (5) | 2 (2.2) | 6 (7) | 0 |
| Cirrhosis | 9 (2) | 6 (3.1) | 3 (1.2) | 0 (0) | 0 | 11 (6) | 4 (4.4) | 7 (8.1) | 0 |
| Immunocompromised | 45 (9) | 14 (7.3) | 22 (8.9) | 9 (16.4) | 0 | 11 (6) | 2 (2.2) | 9 (10.5) | 0 |
| *Site of infection* |  |  |  |  | 3 |  |  |  | 1 |
| Lung | 192 (39) | 106 (55.5) | 74 (30.1) | 12 (21.8) |  | 74 (42) | 42 (48.3) | 32 (37.2) |  |
| Abdomen | 181 (37) | 45 (23.6) | 111 (45.1) | 25 (45.5) |  | 35 (20) | 17 (19.5) | 18 (20.9) |  |
| Urine | 29 (6) | 13 (6.8) | 13 (5.3) | 3 (5.5) |  | 28 (16) | 10 (11.5) | 18 (20.9) |  |
| Primary bacteremia | 10 (2) | 0 (0) | 6 (2.4) | 4 (7.3) |  | 3 (2) | 2 (2.3) | 1 (1.2) |  |
| Neurological | 5 (1) | 3 (1.6) | 2 (0.8) | 0 (0) |  | 4 (2) | 4 (4.6) | 0 (0) |  |
| Soft tissue or line | 26 (5) | 10 (5.2) | 10 (4.1) | 6 (10.9) |  | 6 (3) | 3 (3.5) | 3 (3.5) |  |
| Other | 49 (10) | 14 (7.3) | 30 (12.2) | 5 (9.1) |  | 23 (13) | 9 (10.3) | 14 (16.3) |  |
| SOFA | 8 (6, 9) | 7 (6,8) | 8 (7,10) | 9 (7,11) | 22 | 7 (5, 9) | 6 (4,8) | 8 (5,10) | 16 |
| APACHE II | 25 (21, 31) | 24 (21, 30) | 26 (21, 31) | 27 (22, 30) | 1 | 24 (19, 30) | 24 (18, 29) | 23.5 (20, 30) | 0 |
| Post-surgical admission | 180 (37) | 49 (25.7) | 114 (46.2) | 17 (30.9) | 0 | 26 (15) | 17 (18.9) | 9 (10.5) | 0 |
| **Outcomes** |  |  |  |  | p-value for difference |  |  |  | p-value for difference |
| Mean daily SOFA, mean (SD) |  | 4.93 (2.88) | 6.67 (3.94) | 9.97 (4.60) | <0.001<sup>†</sup> |  |  |  |  |
| 3-month survival, n/N (%) |  | 132/189 (69.8%) | 155/246 (63.0%) | 23/55 (41.8%) | 0.001* |  |  |  |  |
| 28-day survival, n/N (%) |  | 143/190 (75.3%) | 165/247 (66.8%) | 26/55 (47.3%) | <0.001* |  | 72/90 (80.0%) | 57/86 (66.3%) | 0.04* |
| 28-day renal failure-free survival, n/N (%)<sup>1</sup> |  |  |  |  |  |  | 51/75 (68.0%) | 32/60 (53.3%) | 0.08* |
| Renal failure-free days, median (IQR)<sup>2</sup> |  |  |  |  |  |  | 19 (3,26) | 8 (0, 23) | 0.03<sup>‡</sup> |

NYHA= New York Heart Association, COPD=Chronic Obstructive Pulmonary Disease, SOFA=Sequential Organ Failure Assessment, APACHE=Acute Physiology and Chronic Health Evaluation; \* $\chi^2$ test; <sup>†</sup> Kruskal-Wallis test; <sup>‡</sup> Mann-Whitney test; <sup>1</sup> in patients not in renal failure at baseline; <sup>2</sup> in patients who die or experience some renal failure by day-28.

**Table 2.**
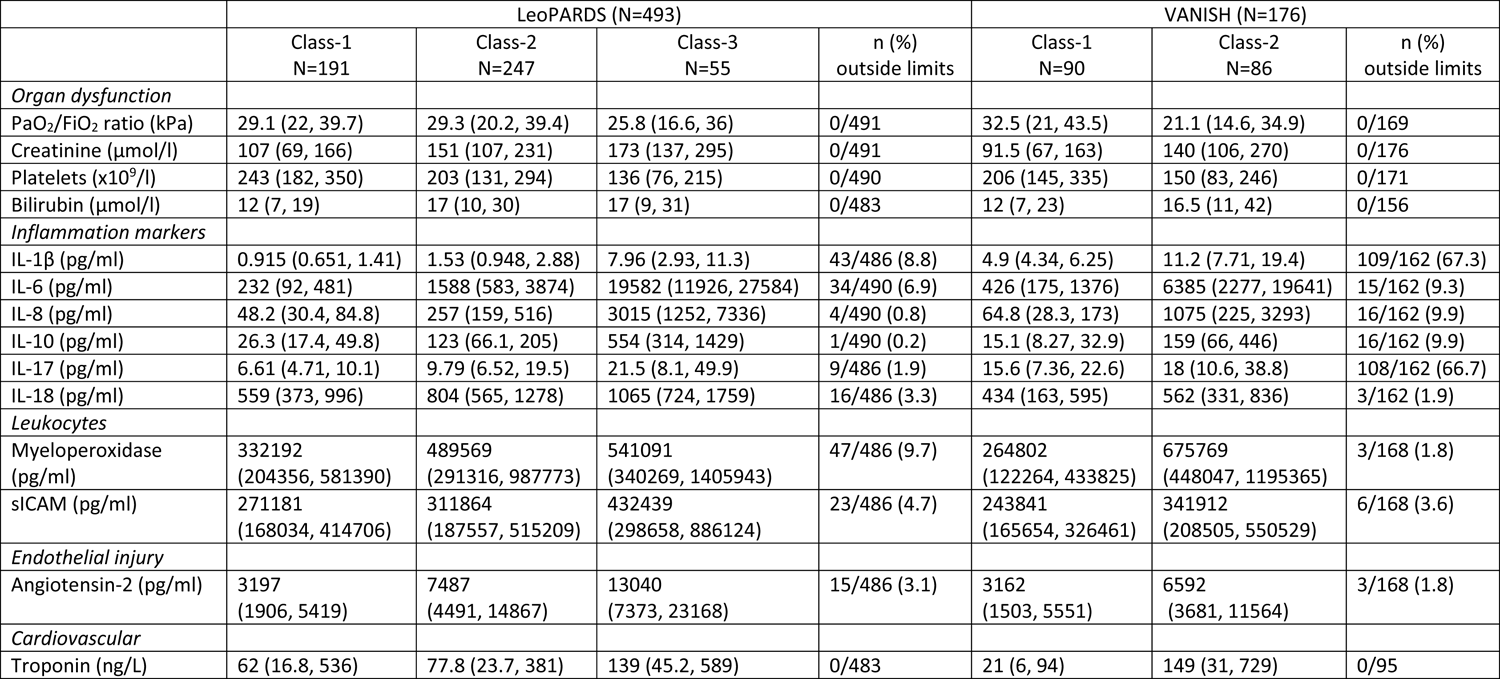
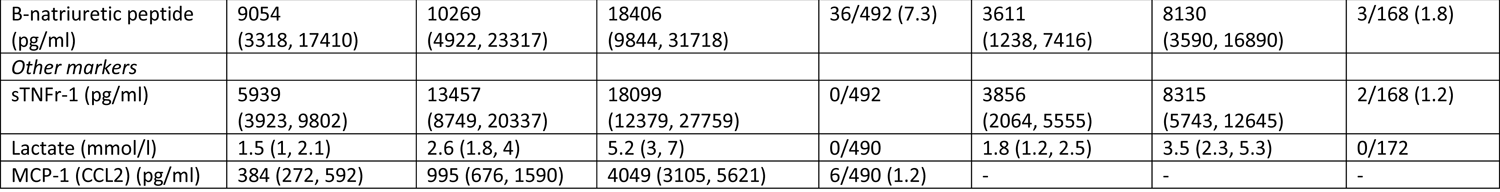
Biomarker data by class, with study participants assigned by highest posterior class probability in LCA. Biomarker values are median (interquartile range) for values within the limit of detection. The number of values outside the limit of detection as a percentage of non-missing values is given.

|  | LeoPARDS (N=493) |  |  |  | VANISH (N=176) |  |  |
| --- | --- | --- | --- | --- | --- | --- | --- |
|  | Class-1<br>N=191 | Class-2<br>N=247 | Class-3<br>N=55 | n (%)<br>outside limits | Class-1<br>N=90 | Class-2<br>N=86 | n (%)<br>outside limits |
| *Organ dysfunction* |  |  |  |  |  |  |  |
| PaO<sub>2</sub>/FiO<sub>2</sub> ratio (kPa) | 29.1 (22, 39.7) | 29.3 (20.2, 39.4) | 25.8 (16.6, 36) | 0/491 | 32.5 (21, 43.5) | 21.1 (14.6, 34.9) | 0/169 |
| Creatinine (μmol/l) | 107 (69, 166) | 151 (107, 231) | 173 (137, 295) | 0/491 | 91.5 (67, 163) | 140 (106, 270) | 0/176 |
| Platelets (x10<sup>9</sup>/l) | 243 (182, 350) | 203 (131, 294) | 136 (76, 215) | 0/490 | 206 (145, 335) | 150 (83, 246) | 0/171 |
| Bilirubin (μmol/l) | 12 (7, 19) | 17 (10, 30) | 17 (9, 31) | 0/483 | 12 (7, 23) | 16.5 (11, 42) | 0/156 |
| *Inflammation markers* |  |  |  |  |  |  |  |
| IL-1β (pg/ml) | 0.915 (0.651, 1.41) | 1.53 (0.948, 2.88) | 7.96 (2.93, 11.3) | 43/486 (8.8) | 4.9 (4.34, 6.25) | 11.2 (7.71, 19.4) | 109/162 (67.3) |
| IL-6 (pg/ml) | 232 (92, 481) | 1588 (583, 3874) | 19582 (11926, 27584) | 34/490 (6.9) | 426 (175, 1376) | 6385 (2277, 19641) | 15/162 (9.3) |
| IL-8 (pg/ml) | 48.2 (30.4, 84.8) | 257 (159, 516) | 3015 (1252, 7336) | 4/490 (0.8) | 64.8 (28.3, 173) | 1075 (225, 3293) | 16/162 (9.9) |
| IL-10 (pg/ml) | 26.3 (17.4, 49.8) | 123 (66.1, 205) | 554 (314, 1429) | 1/490 (0.2) | 15.1 (8.27, 32.9) | 159 (66, 446) | 16/162 (9.9) |
| IL-17 (pg/ml) | 6.61 (4.71, 10.1) | 9.79 (6.52, 19.5) | 21.5 (8.1, 49.9) | 9/486 (1.9) | 15.6 (7.36, 22.6) | 18 (10.6, 38.8) | 108/162 (66.7) |
| IL-18 (pg/ml) | 559 (373, 996) | 804 (565, 1278) | 1065 (724, 1759) | 16/486 (3.3) | 434 (163, 595) | 562 (331, 836) | 3/162 (1.9) |
| *Leukocytes* |  |  |  |  |  |  |  |
| Myeloperoxidase (pg/ml) | 332192 (204356, 581390) | 489569 (291316, 987773) | 541091 (340269, 1405943) | 47/486 (9.7) | 264802 (122264, 433825) | 675769 (448047, 1195365) | 3/168 (1.8) |
| sICAM (pg/ml) | 271181 (168034, 414706) | 311864 (187557, 515209) | 432439 (298658, 886124) | 23/486 (4.7) | 243841 (165654, 326461) | 341912 (208505, 550529) | 6/168 (3.6) |
| *Endothelial injury* |  |  |  |  |  |  |  |
| Angiotensin-2 (pg/ml) | 3197 (1906, 5419) | 7487 (4491, 14867) | 13040 (7373, 23168) | 15/486 (3.1) | 3162 (1503, 5551) | 6592 (3681, 11564) | 3/168 (1.8) |
| *Cardiovascular* |  |  |  |  |  |  |  |
| Troponin (ng/L) | 62 (16.8, 536) | 77.8 (23.7, 381) | 139 (45.2, 589) | 0/483 | 21 (6, 94) | 149 (31, 729) | 0/95 |
| B-natriuretic peptide (pg/ml) | 9054<br>(3318, 17410) | 10269<br>(4922, 23317) | 18406<br>(9844, 31718) | 36/492 (7.3) | 3611<br>(1238, 7416) | 8130<br>(3590, 16890) | 3/168 (1.8) |
| *Other markers* |  |  |  |  |  |  |  |
| sTNFr-1 (pg/ml) | 5939<br>(3923, 9802) | 13457<br>(8749, 20337) | 18099<br>(12379, 27759) | 0/492 | 3856<br>(2064, 5555) | 8315<br>(5743, 12645) | 2/168 (1.2) |
| Lactate (mmol/l) | 1.5 (1, 2.1) | 2.6 (1.8, 4) | 5.2 (3, 7) | 0/490 | 1.8 (1.2, 2.5) | 3.5 (2.3, 5.3) | 0/172 |
| MCP-1 (CCL2) (pg/ml) | 384 (272, 592) | 995 (676, 1590) | 4049 (3105, 5621) | 6/490 (1.2) | - | - | - |

For VANISH, models did not converge when all pre-specified class-predictors were included so reduced models were fit with age, source of infection, APS-APII and post-surgical admission as covariates and the number of censored indicators were reduced by treating values outside the limits as having values equal to the limit, for indicators with fewer than 5 such values. Based on model fit (Table S2, Figure S3) we selected a two-class model with zero covariances between indicators. IL-1β, IL-10, IL-6 and IL-8 showed prominent class separation (Figure 1, Table 2). There was an even split between classes (90 individuals assigned to class 1 and 86 to class 2). Although baseline and demographic characteristics were similar between the two classes, class 1 was associated with greater survival at 28-days and more renal failure free days than class 2 (Table 1). Sensitivity analysis gave comparable results with 97% agreement (Table S4). There was no evidence that treatment effects varied by class for any of the outcomes of the VANISH trial (Figure 2). The effect of vasopressin on renal failure free survival at 28 days compared to norepinephrine was in opposite directions in class 1 (10% reduction in survival (95% CI-31% to 11%)) compared to class 2 (10% increase in survival (95% CI-16% to 35%)) but the confidence intervals were wide (difference in RD 20% (95% CI-13% to 53%)) (Figure 2A). This was also seen for renal failure free days.

### Hierarchical Clustering of Inflammatory Mediators

Since cytokines were the indicators showing greatest separation between LCA classes, we clustered based solely on cytokines using hierarchical clustering analysis (HCA) to understand the impact of clustering approach on sub-phenotype identification and whether clinically meaningful patient clusters could still be identified. While LCA accommodates different data types and handles missing data, HCA is suitable for clustering based on only one type of continuous data and produces clearer inference of how each variable contributes to the clustering and the relation between samples as dendrograms. To include a more comprehensive profiling of cytokines and to allow comparison with previously defined transcriptomic sub-phenotypes (SRS), the GAinS cohort was included in HCA. Both VANISH and GAinS were best described by two classes (Figures 3A and S4) with consensus clustering based on HCA producing highly consistent assignments (94% for VANISH, 100% for GAinS). LeoPARDs could be described by either two or three classes (Figures 3A and S4) with the most consistent group assignment between HCA and consensus clustering being with a three-class model (91%).

**Figure 3.**
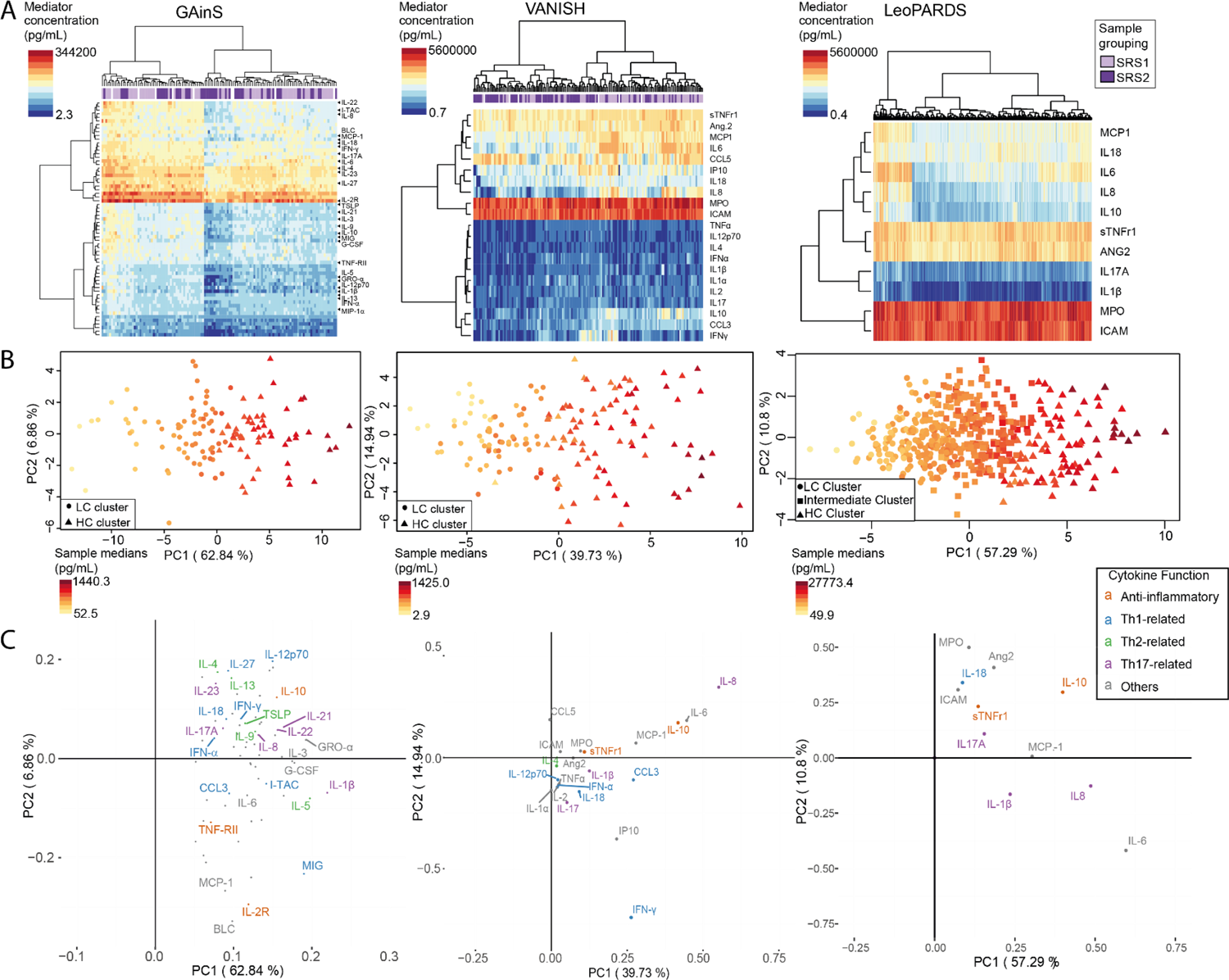
Unsupervised patient structure in GAinS (n=124, left), VANISH (n=155, middle) and LeoPARDS (n=484, right). A) Heatmaps colored by inflammatory mediator concentration, patients shown as columns and mediators as rows, solid bars represent Sepsis Response Signature (SRS) assignments in GAinS and VANISH (light purple=SRS1, dark purple=SRS2, blank=no assignment available). A full-size version of the heatmap in GAinS where all mediators are labelled is available as Figure S5. B) Principal component analysis scores plot where data points are shaded based on median inflammatory mediator concentration by sample (circles= low cytokine cluster (LC), squares,= intermediate cytokine cluster, triangles= high cytokine cluster (HC)). Percentage of variance explained by the principal components (PC) 1/2 are stated in parentheses. C) Loadings of inflammatory mediators on PC1/2. (A, C) For better display in GAinS, we only annotated mediators that are anti-inflammatory, or have been more strongly related with type 1/2/17 T helper cells or have the largest loadings.

**Figure 4.**
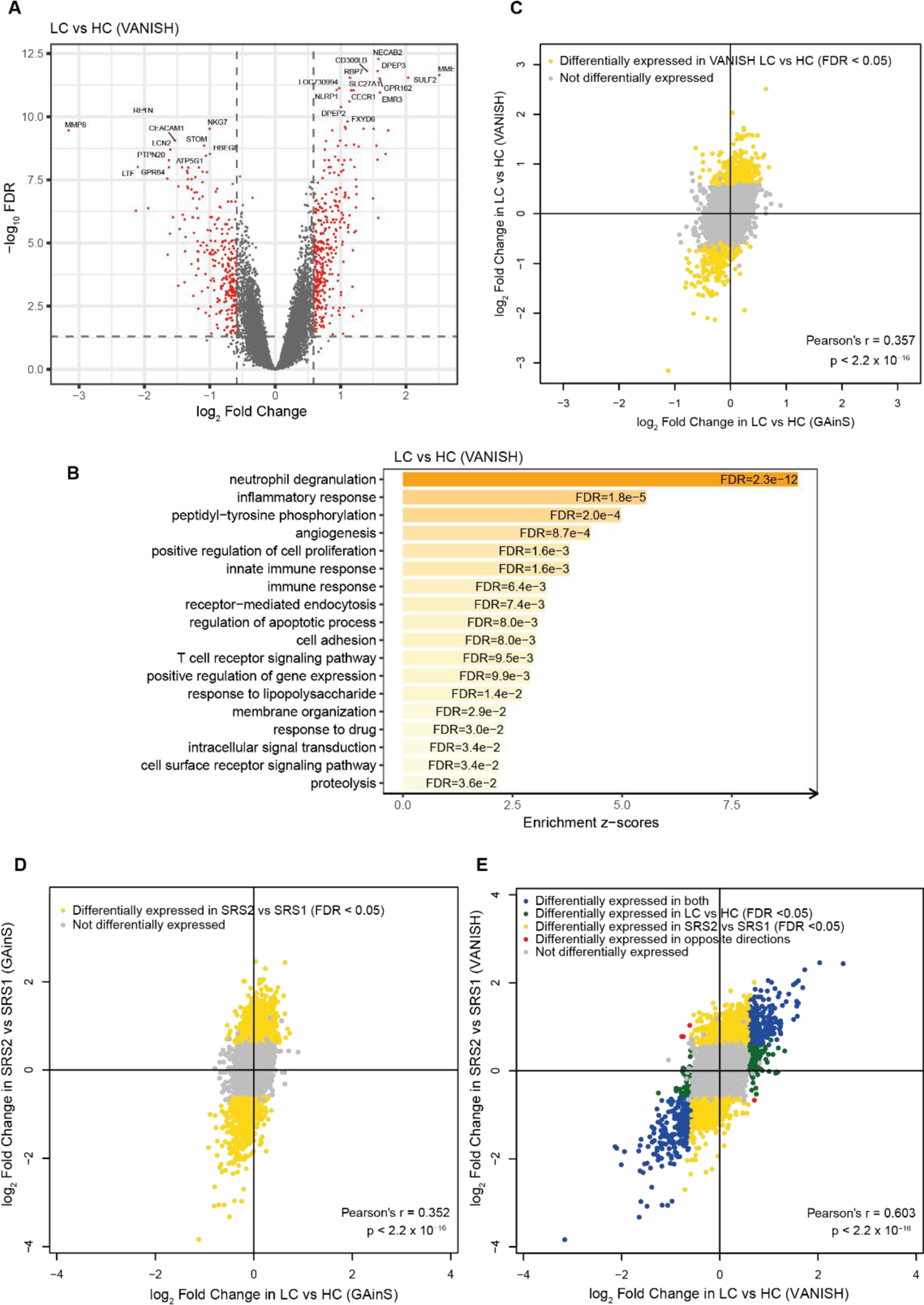
Gene expression comparisons between inflammatory clusters, and correlation between inflammatory cluster comparisons and SRS transcriptomic sub-phenotype comparisons. A) Volcano plot for LC versus HC clusters in VANISH and B) the enriched Gene Ontology Biological Processes of the differentially expressed genes. C) Log2 fold change correlations between the inflammatory cluster comparisons in VANISH and the inflammatory cluster comparisons in GAinS. D, E) Log2 fold change correlations of the inflammatory cluster comparisons and the SRS comparisons in GAinS (D) and in VANISH (E). P-values are shown for tests of correlations using Pearson’s product moment correlation. In the volcano plot (A), red points indicate probes (n=665) for 554 differentially expressed genes (FDR<0.05 and FC>1.5, dashed lines indicate these thresholds). Genes on the right-hand side have higher expression in LC.

Principal component analysis showed separation of the classes along the first component (Figure 1B) which was strongly correlated with the median cytokine concentration of all samples (Spearman’s rho = 0.982/0.904/0.896 for GAinS/VANISH/LeoPARDS). In all cohorts a ‘high cytokine’ (HC) sub-phenotype had higher concentrations of inflammatory mediators than the ‘low cytokine’ (LC) sub-phenotype (Table 3, Table S6 and Figure 1A).

**Table 3.**
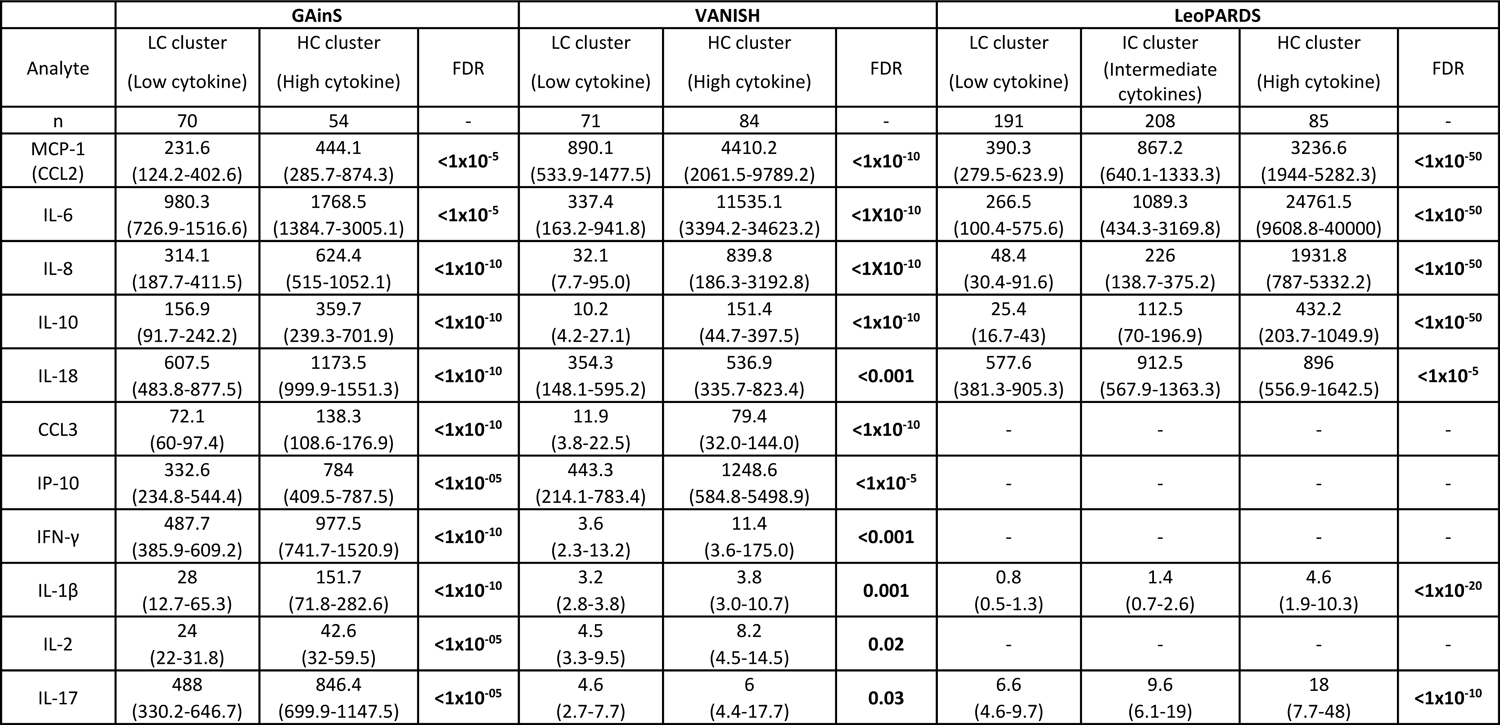
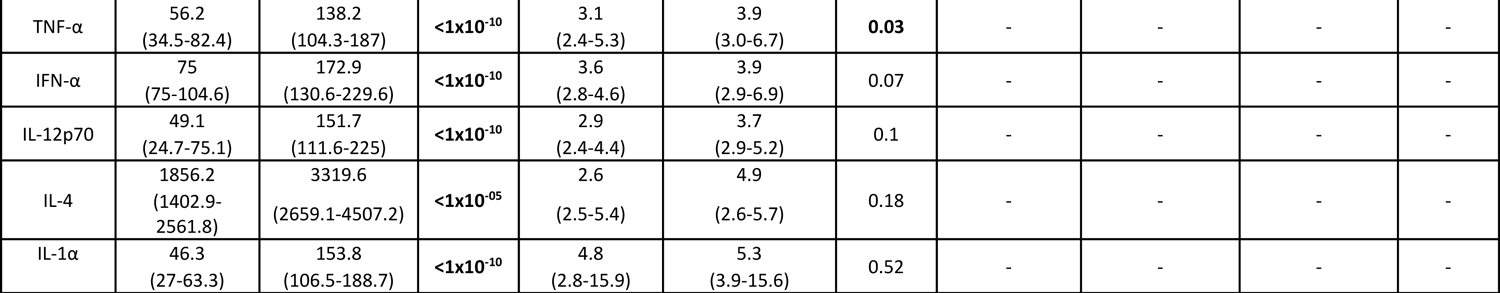
Comparison of inflammatory mediator concentrations between clusters identified in the GAinS, VANISH and LeoPARDS datasets. Only 16 mediators measured in both GAinS and VANISH are shown in this table. The full table including 54 additional mediators are available as supplementary Table S6. Data are given as median and interquartile range in units of pg/mL and comparison has been made with the Mann-Whitney U test for two-group comparisons and the Kruskal-Wallis tests for three-group comparisons. FDR values in bold are those <0.05.

Concordance between LCA and HCA classes was strong. In VANISH 96% of patients in LCA class 2 were also in the HC class and 85% of those in LCA class 1 were in the LC class. In LeoPARDS 98% of patients in LCA class 3 were in the HC class, 88% of those in LCA class 1 were in the LC class and 76% of those in LCA class 2 were in the intermediate HCA class. Patients similarly classified by LCA and HCA had higher LCA class membership probabilities than patients with inconsistent classification (Table S7) suggesting lower certainty of LCA classification in those differently classified.

Clinical differences between HCA sub-phenotypes can be seen in Table 4 and Table S8. Generally, HC sub-phenotype patients showed features of more severe disease such as higher heart rates, lower PaO_2_:FiO_2_ ratio and higher requirement for fluid and norepinephrine. Mortality at 28-days was highest in the HC sub-phenotype in VANISH (p=0.03) and LeoPARDS (p=0.002) with the same trend seen in the GAinS data (Table 4, Table S8). In LeoPARDS poorer survival in the HC sub-phenotype was also seen at 3- and 6-months. Patients in the HC sub-phenotype in VANISH had higher mean total SOFA scores over the duration of their ICU stay than patients in the LC sub-phenotype (p<0.001) as was the case in LeoPARDS where the mean total SOFA increased incrementally from the LC, through the intermediate to HC sub-phenotype (4.1 (2.9-6.1) vs 5.9 (3.9-9) vs 7.3 (4.3-13), p<0.001) (Table S8).

**Table 4.**
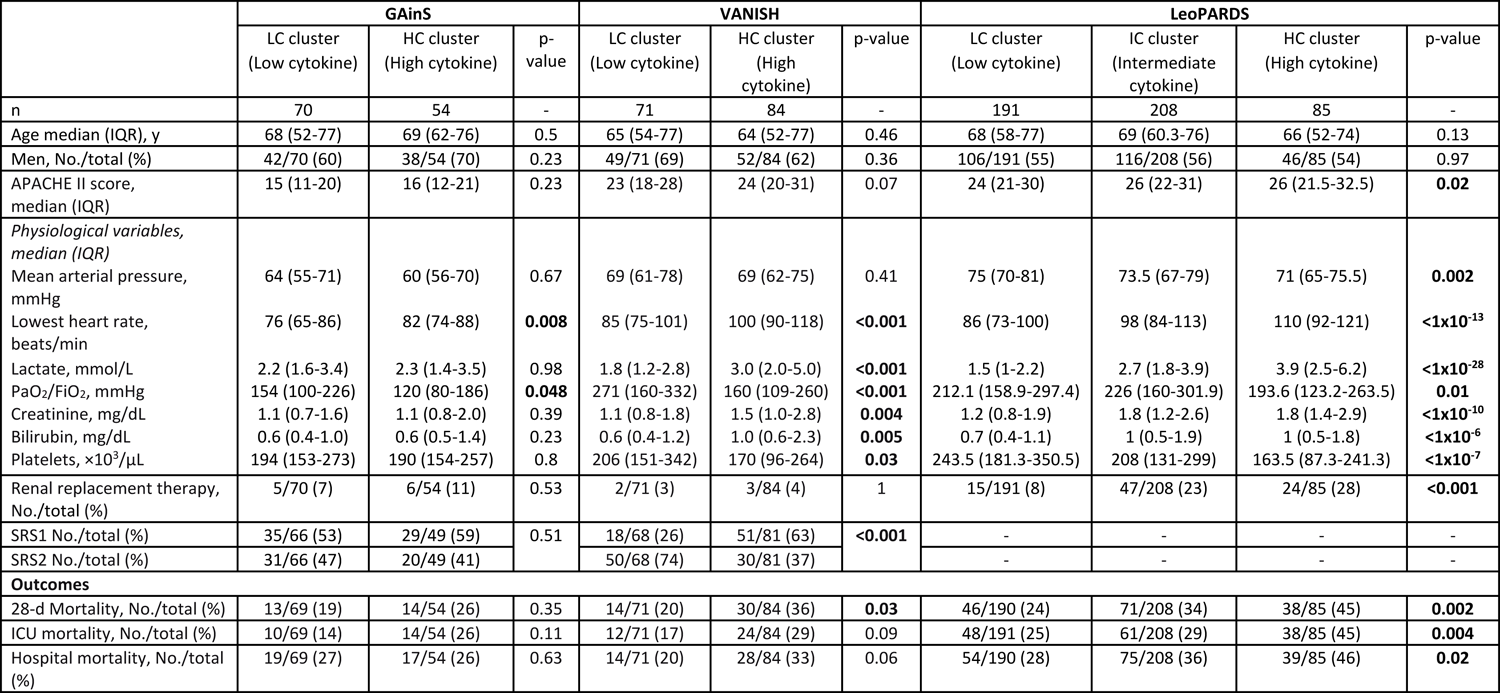
Comparison of baseline variables and clinical outcomes between clusters from the hierarchical cluster analysis in GAinS, VANISH and LeoPARDS. Only a subset of representative variables are shown in this table while the full list of variables compared is available as supplementary Table S8. Continuous variables have been compared with the Mann-Whitney U test or Kruskal-Wallis test and categorical variables with the chi-squared test or Fisher’s exact test (if number of events <10). P-values in bold are those <0.05. No adjustment has been made for multiple comparisons. (APACHE=Acute Physiology and Chronic Health Evaluation, GCS=Glasgow Coma Scale, IQR=interquartile range, ICU=Intensive Care Unit).

|  | GAINs |  |  | VANISH |  |  | LeoPARDS |  |  |  |
| --- | --- | --- | --- | --- | --- | --- | --- | --- | --- | --- |
|  | LC cluster<br>(Low cytokine) | HC cluster<br>(High cytokine) | p-value | LC cluster<br>(Low cytokine) | HC cluster<br>(High cytokine) | p-value | LC cluster<br>(Low cytokine) | IC cluster<br>(Intermediate cytokine) | HC cluster<br>(High cytokine) | p-value |
| n | 70 | 54 | - | 71 | 84 | - | 191 | 208 | 85 | - |
| Age median (IQR), y | 68 (52-77) | 69 (62-76) | 0.5 | 65 (54-77) | 64 (52-77) | 0.46 | 68 (58-77) | 69 (60.3-76) | 66 (52-74) | 0.13 |
| Men, No./total (%) | 42/70 (60) | 38/54 (70) | 0.23 | 49/71 (69) | 52/84 (62) | 0.36 | 106/191 (55) | 116/208 (56) | 46/85 (54) | 0.97 |
| APACHE II score, median (IQR) | 15 (11-20) | 16 (12-21) | 0.23 | 23 (18-28) | 24 (20-31) | 0.07 | 24 (21-30) | 26 (22-31) | 26 (21.5-32.5) | **0.02** |
| *Physiological variables, median (IQR)* |  |  |  |  |  |  |  |  |  |  |
| Mean arterial pressure, mmHg | 64 (55-71) | 60 (56-70) | 0.67 | 69 (61-78) | 69 (62-75) | 0.41 | 75 (70-81) | 73.5 (67-79) | 71 (65-75.5) | **0.002** |
| Lowest heart rate, beats/min | 76 (65-86) | 82 (74-88) | **0.008** | 85 (75-101) | 100 (90-118) | **<0.001** | 86 (73-100) | 98 (84-113) | 110 (92-121) | **<1×10<sup>-13</sup>** |
| Lactate, mmol/L | 2.2 (1.6-3.4) | 2.3 (1.4-3.5) | 0.98 | 1.8 (1.2-2.8) | 3.0 (2.0-5.0) | **<0.001** | 1.5 (1-2.2) | 2.7 (1.8-3.9) | 3.9 (2.5-6.2) | **<1×10<sup>-28</sup>** |
| PaO<sub>2</sub>/FiO<sub>2</sub>, mmHg | 154 (100-226) | 120 (80-186) | **0.048** | 271 (160-332) | 160 (109-260) | **<0.001** | 212.1 (158.9-297.4) | 226 (160-301.9) | 193.6 (123.2-263.5) | **0.01** |
| Creatinine, mg/dL | 1.1 (0.7-1.6) | 1.1 (0.8-2.0) | 0.39 | 1.1 (0.8-1.8) | 1.5 (1.0-2.8) | **0.004** | 1.2 (0.8-1.9) | 1.8 (1.2-2.6) | 1.8 (1.4-2.9) | **<1×10<sup>-10</sup>** |
| Bilirubin, mg/dL | 0.6 (0.4-1.0) | 0.6 (0.5-1.4) | 0.23 | 0.6 (0.4-1.2) | 1.0 (0.6-2.3) | **0.005** | 0.7 (0.4-1.1) | 1 (0.5-1.9) | 1 (0.5-1.8) | **<1×10<sup>-6</sup>** |
| Platelets, ×10<sup>3</sup>/μL | 194 (153-273) | 190 (154-257) | 0.8 | 206 (151-342) | 170 (96-264) | **0.03** | 243.5 (181.3-350.5) | 208 (131-299) | 163.5 (87.3-241.3) | **<1×10<sup>-7</sup>** |
| Renal replacement therapy, No./total (%) | 5/70 (7) | 6/54 (11) | 0.53 | 2/71 (3) | 3/84 (4) | 1 | 15/191 (8) | 47/208 (23) | 24/85 (28) | **<0.001** |
| SRS1 No./total (%) | 35/66 (53) | 29/49 (59) | 0.51 | 18/68 (26) | 51/81 (63) | **<0.001** | - | - | - | - |
| SRS2 No./total (%) | 31/66 (47) | 20/49 (41) |  | 50/68 (74) | 30/81 (37) |  | - | - | - | - |
| **Outcomes** |  |  |  |  |  |  |  |  |  |  |
| 28-d Mortality, No./total (%) | 13/69 (19) | 14/54 (26) | 0.35 | 14/71 (20) | 30/84 (36) | **0.03** | 46/190 (24) | 71/208 (34) | 38/85 (45) | **0.002** |
| ICU mortality, No./total (%) | 10/69 (14) | 14/54 (26) | 0.11 | 12/71 (17) | 24/84 (29) | 0.09 | 48/191 (25) | 61/208 (29) | 38/85 (45) | **0.004** |
| Hospital mortality, No./total (%) | 19/69 (27) | 17/54 (26) | 0.63 | 14/71 (20) | 28/84 (33) | 0.06 | 54/190 (28) | 75/208 (36) | 39/85 (46) | **0.02** |

### Transcriptomic Sub-phenotypes and Inflammatory Clusters

Comparing cytokines between SRS1 and SRS2, significantly different mediators were higher in SRS1 compared to SRS2 (Table S9) with the pro-inflammatory cytokines IL-6, IL-8, MCP-1 and CCL3 being at significantly higher concentrations in SRS1 than SRS2 (FDR<0.05) in both GAinS and VANISH. In the VANISH cohort 63% of HC patients had the SRS1 sub-phenotype, compared with 26% of LC patients (OR of being SRS1 if in the HC vs LC sub-phenotype: 4.72 (95% CI 2.3-9.5), p<0.001, Table 4). Using the LCA classes from VANISH 60% of class 2 patients compared to 35% of class 1 had the SRS1 sub-phenotype (OR of being SRS1 in LCA class 2 vs class 1: 2.8 (95% CI 1.5-5.3), p=0.001). There was no relationship between the HCA clusters and the SRS sub-phenotypes in the GAinS cohort (p=0.51). Dividing patients using both cytokine and SRS sub-phenotypes consistently demonstrated lowest 28-day mortality in LC-SRS2 patients (Figure S6). In GAinS mortality remained predominantly associated with SRS, which was not the case in VANISH.

### Gene Expression Signatures of Inflammatory Clusters

There were no significantly differentially expressed genes between inflammatory sub-phenotypes in GAinS but 554 (665 probes) in VANISH (FDR<0.05 and FC>1.5, Figure 4A), which were enriched for Gene Ontology Biological Process terms involving multiple aspects of the immune and inflammatory response (e.g., ‘neutrophil degranulation’) as well as protein phosphorylation and angiogenesis (Figure 4B). There was a statistically significant, though weak, correlation between the fold changes from the comparisons between inflammatory sub-phenotypes in GAinS and VANISH (Pearson’s r=0.357, p<2.2×10^-16^, Figure 4C) suggesting that the inflammatory sub-phenotypes in both datasets were capturing similar transcriptomic differences, although the magnitudes of difference observed were larger in VANISH.

Significant correlations were observed between differential gene expression in SRS sub-phenotypes and in cytokine clusters (Figure 4D, E), suggesting similarity between the inflammatory mediator clusters and SRS groups at the transcriptome level.

## Discussion

In this study we used LCA of combined clinical and biomarker data in patients with septic shock to derive sepsis inflammatory sub-phenotypes. We then compared these to sub-phenotypes derived using different approaches and data types. Two LCA sub-phenotypes described the patients from the VANISH trial whereas the larger LeoPARDS trial was represented by three. These classifications were broadly consistent with those derived from hierarchical clustering using only cytokine data. Across three data sets a sub-phenotype with greater levels of inflammation was associated with features of worse disease severity and higher mortality. Although some similarities existed between global gene-expression sub-phenotypes and those derived from inflammatory mediators our data suggest that they capture different aspects of sepsis pathology and provide complementary information.

The findings from LCA show similarities to those in ARDS (9–13) where a hyper-inflammatory sub-phenotype is associated with higher levels of inflammatory mediators, high rates of organ failure and death compared to the hypo-inflammatory sub-phenotype. This supports the notion that there are distinct host responses to critical illness or “treatable traits” that are independent of syndromic diagnoses such as ARDS or sepsis (35). This concept is also supported by recent work demonstrating that inflammatory sub-phenotypes can be identified in ventilated patients without ARDS (36) and in pancreatitis (37). The fact that HCA of inflammatory mediators alone provides very similar patient grouping to LCA of clinical and inflammatory data suggests that most group separation derives from inflammatory mediators that are not routinely measured in clinical practice and provides evidence that these groups are robust to the choice of clustering approach, enhancing their validity. Previous clustering of inflammatory mediators in sepsis has identified differing numbers of sub-phenotypes (15–19). Differences in patient populations, number of subjects and mediators measured make comparisons difficult. Yet, in common with our work, clusters with high cytokine concentrations have been identified amongst patients with sepsis that are associated with shock, organ dysfunction and increased mortality (15, 18).

ARDS sub-phenotypes have shown potential to identify patients who may benefit from specific treatments (10, 12, 13). However, here we found no sub-phenotype treatment interaction in sepsis. This may reflect the specific treatments used in these trials, or that they were not powered for identifying differential responses. We have previously demonstrated an interaction between SRS gene expression sub-phenotypes and use of hydrocortisone (22), with patients in the SRS2, low mortality, group having higher mortality if given hydrocortisone compared with placebo, so it is noteworthy that no differential effect was seen using LCA inflammatory sub-phenotypes for this treatment.

Our data suggests that inflammatory and gene-expression sub-phenotypes are not comparable, although there was some overlap. Although VANISH patients in the high inflammation groups were more likely to also be in the SRS1 transcriptomic sub-phenotype, a third of patients were not and in GAinS we found no significant relationship. This incomplete overlap between cytokine and transcriptomic sub-phenotypes could explain the differences seen in the interaction between treatment and sub-phenotype described here and previously (22). Differential gene expression analysis was able to highlight biological processes that distinguished the high cytokine from low cytokine sub-phenotypes, such as neutrophil degranulation and the inflammatory response. It is of note that it was generic pathways and not individual cytokines that were differentially expressed. This potentially explains the globally higher cytokine levels instead of differences in individual cytokines seen in the high cytokine sub-phenotypes.

Comparison of cytokines between SRS sub-phenotypes also provided support for an overlap of these groups and cytokine groups. IL-6, MCP-1, IL-8 and CCL3 were significantly higher in SRS1 than SRS2 in both GAinS and VANISH while eight other markers (IL-10, IP-10, IL-2R, IL-31, HGF, MIP-3α, MMP-1 and Ang-2) were also significantly higher in SRS1 in one of the two cohorts (six of these were only available in one of the two cohorts). Gene expression signatures on SRS discovery indicated that SRS1 is characterized by T-cell exhaustion, endotoxin tolerance and HLA class II down regulation (20). Both excessive pro-inflammatory cytokines (IL-6, IL-27, IFN-α) and anti-inflammatory cytokines (IL-10) can lead to T cell exhaustion (38–40) and IL-1β, TNFα, and IL-10 can induce cytokine-mediated tolerance of monocytes (40–43). In our study, other than IL-6, SRS1 had statistically higher concentrations of IL-10 in the VANISH cohort but not GAinS, although the trend was in the same direction. In GAinS, IFN-α and TNFα levels were higher in SRS1 compared with SRS2 although not reaching statistical significance. Further work is needed to understand the causal and temporal relationship between inflammation and transition into the SRS1 sub-phenotype with recent evidence, for example, of granulopoietic dysfunction involving specific neutrophil subsets (44).

There are limitations to this work. Firstly, because of differences in patient cohorts, sample sizes, analytical platforms and panels of mediators measured it is impossible to be certain that the sub-phenotypes described are the same between patient cohorts. These differences may account for some of the discrepancy in clinical differences between cytokine clusters in GAinS, VANISH and LeoPARDS. Similarly, different timing of sample collection in relationship to the onset of the inflammatory response and sepsis complicates interpretation of the relationship between cytokine and transcriptomic sub-phenotypes across the datasets. However, the similarities in patterns of clinical features and cytokine levels between the clusters, irrespective of patient cohorts is reassuring. There were also limitations associated with the platforms used to measure the cytokines. The multiplex system used in VANISH resulted in several mediators with concentrations below the level of quantification. Censoring of this data with the value of the lower limit will have introduced imprecision, compromising the interpretation of the importance of these mediators. Finally, none of the trial datasets were designed with the intention of performing a sub-group analysis as reported here and as such are likely to be underpowered to detect a sub-group/treatment interaction. Therefore, it is important that these results are viewed as exploratory and not as conclusive evidence that no sub-phenotype treatment interaction exists.

In conclusion, we found that both LCA and HCA identified a sub-phenotype of patients with high levels of inflammatory mediators and worse disease severity and outcomes. Inflammatory sub-phenotypes showed some similarities with transcriptomic sub-phenotypes although these two approaches did not identify identical patient groups. We conclude that future patient stratification, either for prognostication or for treatment allocation, will need to combine multiple types of biological data, such as gene expression and cytokine data, to allow the most accurate classification and that the balance of data types may depend on the clinical question that needs to be addressed.

## Funding

This paper presents independent research funded by the UK National Institute for Health and Care Research (NIHR) under its Research for Patient Benefit program (Grant number PB-PG-0610-22350), an NIHR Clinician Scientist Award (NIHR/CS/009/007), and an NIHR Research Professor award (RP-2015-06-018) held by Prof Gordon and the U.K. Efficacy and Mechanism Evaluation (EME) Programme, a Medical Research Council (MRC) and NIHR partnership (16/33/01; 08/99/08; 11/14/08). It was also supported by the NIHR Imperial Biomedical Research Centre, and also by the UK Intensive Care Foundation. Further support was received from Wellcome Trust Grant (090532/Z/09/Z) to core facilities at the Wellcome Centre for Human Genetics, a Wellcome Trust Investigator Award (204969/Z/16/Z) to Prof Knight, the Chinese Academy of Medical Sciences (CAMS) Innovation Fund for Medical Science (CIFMS), China (grant number: 2018-I2M-2-002), the NIHR Oxford Biomedical Research Centre, and from a China Scholarship Council – University of Oxford Scholarship held by Mi. Drs Davenport and Burnham are supported by Wellcome Sanger Institute Core Funding from the Wellcome Trust [206194 and 108413/A/15/D]. For the purpose of Open Access, the authors have applied a CC BY public copyright license to any Author Accepted Manuscript version arising from this submission. The authors Dr Antcliffe and Prof Gordon are affiliated with the Department of Health and Social Care, Centre for Antimicrobial Optimization at Imperial College, London. The views expressed in this article are those of the authors and not necessarily those of the MRC, National Health Service, NIHR, or Department of Health. The funders of the study had no role in design and conduct of the study; collection, management, analysis, and interpretation of the data; and preparation, review, or approval of the manuscript, or the decision to submit for publication.

## Author contributions

Professors Gordon, Knight and Shankar-Hari had full access to all of the data in the study, take responsibility for the integrity of the data and the accuracy of the data analysis and had final responsibility for the final decision to submit for publication.

*Study concept and design:* DBA, YM, KLB, TP, COK, DFM, JCK, ACG, MS-H

*Acquisition, analysis, or interpretation of data:* All authors

*Drafting of the manuscript*: DBA, YM, SS, MS-H

*Critical revision of the manuscript for important intellectual content:* All authors

*Statistical Analysis:* DBA, YM, SS, TP, MS-H

*Obtained funding:* ACG, JCK, MS-H, TP, COK, DFM

*Administrative, technical, or material support:* DBA, YM, KLB, FA-B, JCK, ACG, MS-H

All authors read the final draft of the manuscript and confirm to the accuracy / integrity of the work.

## Competing interests

ACG reports that he has received speaker fees from Orion Corporation Orion Pharma and Amomed Pharma. He has consulted for Ferring Pharmaceuticals, Tenax Therapeutics, and received grant support from Orion Corporation Orion Pharma, and Tenax Therapeutics with funds paid to his institution. DFM reports grants to his institution from the MRC/NIHR EME programme for the conduct of this work. Outside the submitted work, DFM reports personal fees for consultancy from Bayer, GSK, Boehringer Ingelheim, Eli Lilly, Novartis and SOBI and for being a member of the data monitoring and ethics committee for Vir Biotechnology and Faron studies and as an educational seminar speaker for GSK. DFM has received funding to his institution from the NIHR, MRC, Wellcome Trust, Innovate UK, Northern Ireland Health and Social Care Research and Development Office, Novavax and Randox. In addition, DFM is a named inventor on a patent US8962032 covering the use of sialic acid–bearing nanoparticles as anti-inflammatory agents issued to his institution. DFM was a Director of Research for the Intensive Care Society and is the Director for the MRC/NIHR EME Programme.

MSH is a Director of Research for the Intensive Care Society, Member of the MRC/NIHR EME Programme, UK Representative of the ESICM, and on the Board of International Sepsis Forum. MSH declares that he has done advisory board activity either directly or indirectly through International Sepsis Forum for Biotest, Endpoint Health, Janssen, Pfizer, and Santersus, with payments going into the unrestricted institutional research funds.

CO reports grants to her institution from the MRC/NIHR EME programme for the conduct of this work. Outside the submitted work, CO reports personal fees for consultancy from INSMED and for committee membership for the California Institute of Regenerative Medicine. Outside of the submitted work CO has received research funding from MRC, Wellcome Trust, NIHSC R&D and Innovate UK. CO’s spouse has received personal fees for consultancy outside of the submitted work from Bayer, GSK, Boehringer Ingelheim, Eli Lilly, Novartis and SOBI and fees for being a member of the data monitoring and ethics committee for Vir Biotechnology and Faron studies and as an educational seminar speaker for GSK. CO’s spouse is a named inventor on a patent US8962032 covering the use of sialic acid–bearing nanoparticles as anti-inflammatory agents issued to his institution.

All other authors declare no conflict of interest directly applicable to this research.

## Data availability

The gene expression data is available on ArrayExpress (accession: E-MTAB-4421 / E-MTAB-4451 / E-MTAB-5273 / E-MTAB-5274 / E-MTAB-7581). Individual participant data that underlie the results in this article, after de-identification (text, table, and figures) and inflammatory mediator data will be made available from the corresponding author on submission of a data request application.

## Data Availability

Gene-expression data is available on ArrayExpress. Individual participant data that underlie the results in this article, after de-identification (text, table, and figures) and inflammatory mediator data will be made available from the corresponding author on submission of a data request application.

https://www.ebi.ac.uk/arrayexpress/

## Notes

### Author Declarations

Ethical approval was obtained from the following ethics committee for aspects of this work: Oxford A research ethics committee ref 12/SC/0014 (VANISH) London-Harrow Research and Ethics Committee ref 13/LO/0365 (LeoPARDS) Scotland A Research Ethics Committee ref 05/MRE00/38 and Berkshire Research Ethics Committee ref 08/H0505/78 (GAinS) all gave ethical approval for this work.

