## Supplemental data for "Inflammatory sub-phenotypes in sepsis: relationship to outcomes, treatment effect and transcriptomic sub-phenotypes"

### Supplementary Methods

**Table S1:** Mediators used for hierarchical cluster analysis in the three datasets

**Table S2:** Fit statistics for latent class models

**Table S3:** Estimated class distribution, indicator means and separation for the LeoPARDS trial.

**Table S4:** Differences in class assignment for main and sensitivity analysis

**Table S5:** Model coefficients from multinomial regression model

**Table S6.** Comparison of inflammatory mediator concentrations between hierarchical clusters identified in the GAinS, VANISH and LeoPARDS datasets

**Table S7:** Comparison of the LCA probabilities of class membership between those patients similarly classified by LCA and HCA and those that were not

**Table S8:** Comparison of baseline variables and clinical outcomes between clusters from the hierarchical cluster analysis models in GAinS, VANISH and LeoPARDS

**Table S9:** Differential cytokine abundance analysis between the SRS1 and SRS2 transcriptomic sub-phenotypes in GAinS and VANISH

**Figure S1:** Plots of LCA model fit indicators in the LeoPARDS trial

**Figure S2:** Class-specific sensitivity, specificity and c-statistics for multinomial logit models with increasing number of predictors, LeoPARDS trial

**Figure S3:** Plots of LCA model fit indicators in the VANISH trial

**Figure S4:** Assessment of hierarchical clustering performance in full cytokine panels of GAinS, VANISH and LeoPARDS

**Figure S5:** Hierarchical clustering of inflammatory mediators in baseline samples in GAinS

**Figure S6.** Comparison of 28-day mortality of patients in GAinS and VANISH stratified by SRS, cytokine clusters and a combination of the sub-phenotypes

**Supplementary References**

**Supplementary Methods**

***The Vasopressin vs Norepinephrine as Initial Therapy in Septic Shock (VANISH) Trial***

*Design*

VANISH was a double-blind, factorial (2x2), randomized controlled trial conducted in 18 intensive care units in the United Kingdom.

*Treatment Regimen*

Patients were randomized to receive blinded vasopressin (0-0·06 U/min) or norepinephrine (0-12 μg/min) as first line vasopressor to maintain the mean arterial blood pressure after adequate fluid resuscitation within the first 6 hours after onset of septic shock. If the maximal dose of the first study drug was reached patients were randomized to receive either hydrocortisone or placebo, the second study drug. If the patient was still hypotensive after the first dose of study drug 2 then open label catecholamine vasopressors could be used. If the patient was still not responding to open label vasopressors, then open label IV hydrocortisone could be given as a rescue therapy.

Blood sampling occurred in 10 centers where research nurses were available. The baseline sample was collected up to 12h after study drug 1 was administered, however, the majority were collected at or prior to drug administration.

*Patients*

*Inclusion Criteria*

Adult patients who required vasopressors for the management of sepsis despite adequate fluid resuscitation.

Patients needed to fulfil the following inclusion criteria:

- Fulfil 2/4 of the systemic inflammatory response syndrome criteria (1) due to known or suspected infection within the previous 24 hours.
- Hypotension despite adequate intravenous fluid resuscitation.

*Exclusion Criteria*

Patients were excluded if any of the following criteria were met:

- The patient had received a continuous infusion of vasopressors previously during the ICU admission (other than vasopressors used as emergency treatment [for less than six hours] to stabilize the patient during this episode). Vasopressors included norepinephrine, epinephrine, vasopressin, dopamine, metaraminol, phenylephrine, and (intermittent) terlipressin.
- Regular systemic corticosteroid therapy within the previous three months (this did not include inhaled steroid therapy).
- Known adrenal dysfunction / insufficiency.
- End-stage renal failure (i.e. requiring long term dialysis)

- The physician and team were not committed to full active care.
- The patient was known to be pregnant.
- The patient had known acute mesenteric ischemia.
- The patient was known to have Raynaud's phenomenon, systemic sclerosis or other vasospastic diseases.
- The patient had been enrolled in another clinical trial of an investigational medicinal product within 30 days or was enrolled in another interventional study that might interact with the study drugs.
- The patient had a history of anaphylaxis or hypersensitivity to any study drug.

*Informed Consent*

Due to the emergency nature of the trial, a waiver of initial consent was granted. Patients could be enrolled without prospective consent and then written consent was obtained from the patient or a personal or professional legal representative as soon as possible. For cases in which a legal representative gave consent, retrospective written consent was sought once the patient regained decision-making capacity.

*Blood Sampling*

Plasma was collected on the first day of septic shock. Samples were separated locally, frozen, and sent to the coordinating center in batches for storage and subsequent analysis.

*Patient Selection for Current Study*

One patient was excluded from the VANISH cohort due to uncertainty about the timing of the baseline blood sample and another because they had been incorrectly enrolled into the trial and had subsequently been excluded, so had no clinical data. For hierarchical clustering all patients were included where there was a complete set of baseline cytokine data as this method is unable to handle missing data. Missing cytokine data were due to technical issues so were considered missing at random.

***The Levosimendan for the Prevention of Acute Organ Dysfunction in Sepsis (LeoPARDS) Trial***

*Design*

LeoPARDS was a multicenter double-blind, placebo-controlled trial conducted in 34 Intensive Care Units (ICUs) in the United Kingdom between January 2014 and December 2015.

*Treatment Regimen*

Patients received levosimendan or placebo for 24 hours in addition to standard care. Drug infusion was started at 0.1 µg/kg per minute and increased after 2-4 hours to 0.2 µg/kg per minute for the remainder of the 24 hours. All other aspects of clinical care were at the discretion of the treating clinicians. Plasma samples were collected prior to randomization on the day of inclusion (day 1).

*Patients*

*Inclusion Criteria*

Adult patients with septic shock who required vasopressors for at least 4 hours and were recruited within 24 hours of meeting inclusion criteria were eligible for entry into the study. Inclusion criteria used the internationally established consensus definitions of sepsis at that time (2).

• Fulfil 2/4 of the criteria of the systemic inflammatory response syndrome (SIRS) due to known or suspected infection within the previous 24 hours.

The SIRS criteria were:

(1) fever (>38 °C) or hypothermia (< 36 °C),

(2) tachycardia (heart rate > 90 beats per minute),

(3) tachypnoea (respiratory rate > 20 breaths per minute or PaCO2 < 4.3 kPa) or need for mechanical ventilation,

(4) abnormal leukocyte count (> 12,000 cells/mm3, < 4000 cells/mm3, or > 10 immature [band] forms).

• Hypotension, despite adequate intravenous fluid resuscitation, requiring treatment with a vasopressor infusion (e.g. norepinephrine / epinephrine / vasopressin analogue) for at least four hours and still having an ongoing vasopressor requirement at the time of randomization. Adequate fluid resuscitation was achieved using repeated fluid challenges.

*Exclusion Criteria*

• More than 24 hours since meeting all the inclusion criteria

• End-stage renal failure at presentation (previously dialysis-dependent)

• Severe chronic hepatic impairment (Child-Pugh class C)

• A history of Torsades de Pointes

• Known significant mechanical obstructions affecting ventricular filling or outflow or both.

• Treatment limitation decision in place (e.g, Do Not Attempt Resuscitation or not for ventilation/ dialysis)

• Known or estimated weight >135kg

• Known to be pregnant

• Previous treatment with levosimendan within 30 days

• Known hypersensitivity to levosimendan or any of the excipients

• Known to have received another investigational medicinal product within 30 days or currently in another interventional trial that might interact with the study drug.

*Blood Sampling*

Plasma was collected on the first day of septic shock. Samples were separated locally, frozen, and sent to the coordinating center in batches for storage and subsequent analysis.

### ***Genomic Advances in Sepsis (GAinS)***

*Design*

Adult patients (>18y) were recruited at 23 UK ICUs between 2006 and 2015 as part of the UK Genomic Advances in Sepsis (GAinS) study (ukccggains.com) (3, 4). Ethics approval was granted nationally (REC Reference Number 05/MRE00/38 and 08/H0505/78) and for individual participating centers, with informed consent obtained from patients or their legal representative. Sepsis was diagnosed according to ACCP/SCCM guidelines, and all patients showed evidence of organ dysfunction (4). Community acquired pneumonia (CAP) was defined as a febrile illness associated with a cough, sputum production, breathlessness, leukocytosis and radiological features of pneumonia which was acquired in the community or within two days of ICU admission (5, 6). Fecal peritonitis (FP) was diagnosed at laparotomy as inflammation of the peritoneal membrane secondary to large bowel perforation and fecal contamination (7). Demographics and clinical covariates were recorded using an electronic case report form which included details of the results of microbiological investigations as previously described (4).

*Exclusion Criteria*

Patient or legal representative unwilling or unable to give consent; age <18 years; pregnancy; advanced directive to withhold or withdraw life sustaining treatment; admission for palliative care only; or immune-compromise.

*Blood Sampling*

Plasma was collected on the first day of ICU admission. Samples were separated locally, frozen, and sent to the coordinating center in batches for storage and subsequent analysis.

*Patient Selection for Current Study*

From the GAinS cohort, 96 CAP patients and 32 FP patients were included for cytokine profiling in this study. Four day 1 CAP samples failed quality control thus were excluded in the subsequent clustering analysis of 124 patients in total.

***Biomarker Measurement***

Plasma was collected on the first day of ICU admission (GAinS) or of septic shock (VANISH and LeoPARDS).

Biomarkers not collected as part of the clinical data were measured using a combination of LEGENDplex™ multiplex assays (BioLegend, San Diego, USA), enzyme linked immunosorbent assay (ELISA) ELLA™ multiplex assays (ProteinSimple, San Jose, CA, USA), ELLA™ Simple Plex assays (ProteinSimple, San Jose, CA, USA), Duoset ELISA (R&D Systems, Minneapolis, MN,

USA) assays, sandwich ELISA assays (Abcam, Cambridge, UK) and the ProcartaPlex™ Luminex platform (ThermoFischer Scientific, Waltham, MA, USA).

***Latent Class Analysis***

Latent class analysis (LCA) is used to estimate a latent (i.e. unobserved) categorical variable which assigns individuals to groups (*classes*), when we have a set of observed data (*indicators*) which we believe is distributed differently for each class. LCA is a type of finite mixture model that jointly estimates a model for each of the indicators, with each indicator distribution being a mixture of class-specific distributions. Simultaneously, a multinomial logistic model for probabilities of class membership is estimated. The number of classes is specified in the model, but models with different numbers of classes can be compared. We used LCA to identify latent sub-phenotypes in adults with septic shock based on observed biomarker data.

LCA analyses were carried out separately for LeoPARDS and VANISH cohorts, agnostic of outcome, as both had the same biomarkers and clinical variables available. Clinical and inflammatory biomarkers were chosen for inclusion in the LCA models based on their associations with sepsis pathophysiology ($PaO_2/FiO_2$ ratio, creatinine, platelets, bilirubin, IL-1β, IL-6, IL-8, IL-10, IL-17, IL-18, MPO, sICAM, ANG-2, troponin, NT-proBNP, sTNFr, lactate and MCP-1 (CCL2)). The acute physiology element of the APACHE II score (APS-AP II) was included as a covariate in the indicator models as we expected APS-AP II to be associated with the biomarkers independently of subclass. Other baseline clinical and demographic variables (age, ethnicity, BMI, co-morbidities (any of NYHA class IV, severe COPD, chronic renal failure, cirrhosis, immunodeficiency), site of infection (lung/abdomen/urine/other),

SOFA score, the acute physiology element of the APACHE II score (APS-AP II), and post-surgical admission) which may be predictive of subclass were included in the model as class predictors.

Histograms and pairwise correlations were used to assess distributions, outliers, and skewness (highly likely for the cytokine data). For assay data, the number and percentage of values below or above the limits of detection were recorded. Normal distributions were used for the continuous indicators, applying natural log transformations as necessary. Observations above or below the limits of detection were included in the analysis but treated as censored. All variables were standardized to have a mean of 0 and standard deviation of 1, with parameters taken from the data within the limits of detection. The number and proportion of missing observations were described for all biomarkers, clinical variables and demographic characteristics. If a patient has any missing individual indicators, LCA still allows the rest of the complete data to be included, implicitly making the assumption that the data are missing at random (i.e. the probability of missingness depends only on the observed data and not any missing data). This is reasonable for the biomarker data as missing individual indicators are likely to be due to technical issues.

LCA models were fit in three stages. First conditional independence was assumed (all covariances constrained to zero) and no covariates predicting class membership were included. Secondly, pre-specified clinical and demographic variables measured at baseline were included as covariates predicting class membership. Thirdly, variance assumptions concerning indicators were relaxed to allow (a) non-constant residual variance across classes (b) non-zero covariances (c) both of these. It was not possible within the software used to model covariances between censored variables. For each stage we first fit a 1 –class model, then increased the number of classes by 1 until convergence could not be achieved. A number of strategies were used to achieve convergence, namely: (i) for a k-class model, using starting values from a k-1 class model (ii) using alternative integration methods (iii) reducing the number of censored indicators by treating values outside the limits as having values equal to the limit, for indicators with less than 5 such values (iv) reducing the number of class predictors, selecting covariates which improved model fit based on likelihood ratio tests.

For each LCA model, the class means were estimated and differences across classes compared to determine which indicators showed the most separation across classes. For each model and each participant, the probability of an individual being in each class was predicted, with the probabilities for a participant summing to 1 across the classes. Each participant could then be assigned to the class with which they have the highest class probability. A series of latent class models were fit, increasing the number of classes until convergence could no longer be achieved. As many of the biomarkers are correlated we fitted models allowing for correlation between continuous indicators. Simpler models with zero correlation were selected if these gave a similar or better fit. The Bayesian Information Criterion (BIC) was the primary measure of model selection, with smaller values indicating better fit. We also considered the Akaike Information Criterion (AIC), log likelihood, entropy (a measure of class separation between 0 and 1) (8), class sizes (with very small classes being indicative of overfitting) and the mean probability of class assignment, averaged over participants in the class. We also assessed the class means and sizes to see if the substantive interpretation of the classes differed across models and plots of the change in fit statistics with the number of classes were used to determine where additional classes gave limited improvement in fit (9).

*Sensitivity analysis*

In the main analysis we drew a distinction between class-defining variables (Indicators) and class-predicting variables. As a sensitivity analysis we compared the class groupings when including all variables as indicators in the latent class model, following earlier work by Calfee and colleagues (10–14). As LeoPARDS was the larger study with more data, in this dataset, we also constructed a model to predict latent class using a reduced set of indicators. A series of multinomial logit models were estimated with latent class as the outcome and an increasing number of biomarkers as predictors, added in the order of greatest separation between classes. The probability of being in each class was predicted for each patient, and patients were assigned to the class with the highest probability. The class-specific sensitivity, specificity and c-statistics for each model were calculated by comparing the "gold standard" class of the latent class model with the "test" class of the multinomial model. The final number of markers was chosen as the model for which the addition of further variables would bring negligible increases in accuracy.

*Clinical Outcomes*

The primary outcome in this analysis, for the data from the LeoPARDS trial, was survival at 3 months. Mean total SOFA score over 28 days (or ICU stay, whichever was shorter) and survival to 28 days were examined as secondary outcomes. For the VANISH trial, we examined survival to 28 days, as survival to 3 months was not available. Survival free of renal failure to 28 days amongst patients not in renal failure at baseline, and days alive and free of renal failure up to 28 days for all other patients (those who died or experienced some renal failure by day 28) were also examined.

Treatment effect was assessed primarily on an intention to treat basis, with the exception of hydrocortisone vs. placebo, in the VANISH trial as patients were only eligible to receive hydrocortisone/placebo if they reached the maximum infusion of the first study drug. As there was no interaction between the study drugs, and given the limited power of the analysis, only patients eligible to receive the second drug were included for this comparison. Treatment effects were expressed as a risk difference (RD), and the difference in treatment effects across classes as the difference in RD. 95% confidence intervals for the RD and difference in RD were calculated using linear regression with robust standard errors (15). For mean total SOFA, we presented the mean and standard deviation (SD), with differences between classes or treatment arms expressed as a difference in means. As mean total SOFA was skewed, 95% confidence intervals were calculated with bootstrapping, as was done in the main trial analysis. The median and interquartile range was presented for days alive and free of renal failure, again with bootstrap confidence intervals. For continuous variables, permutation tests were used to calculate p-values for the treatment-class interaction.

***Hierarchical Clustering Based on Inflammatory Mediator Concentrations***

As a comparison to LCA defined sub-phenotypes, hierarchical cluster analysis (HCA) was applied to three datasets independently on the full panels of inflammatory mediators. Only patients with complete baseline mediator panels were used for hierarchical cluster analysis (HCA). Mediator concentrations that fell outside of the limits of quantification were censored at the quantification limits (Table S1). All data were natural log transformed prior to clustering. Dissimilarity between samples was measured by Euclidean distance. Ward's method was used as linkage for cluster agglomeration. The optimal number of clusters was defined by inspection of the dendrograms, comparison of test sample distances to cluster centers in cross-validation, and by determining cluster robustness by consensus clustering (16) (Figure S4).

All clustering was performed in R (17).

#### ***Differential Gene Expression***

Microarray data from GAinS and VANISH was co-normalized using the vsn package (18) and batch corrected using the ComBat function from the sva package (19) in R, resulting in 28220 communal probes after quality control. Gene expression data were available at baseline for 115 patients in GAinS and 149 in VANISH following additional quality control with MixupMapper (20). Differentially expressed genes between sub-phenotypes were identified using the limma package (21), which fits a linear model to the expression of each gene and applies an empirical Bayes smoothing to the standard errors of the estimated log-fold changes to account for the overall variance. Pathway enrichment analysis was performed with the R package XGR (22), using annotations of Gene Ontology Biological Process. The differentially expressed genes were tested against the background of all genes with a hypergeometric distribution. Contrasts between the cytokine clusters or SRS groups were limited to the same subsets of patients with both assignments available.

#### ***Statistical Analysis***

Correlation was evaluated with either Pearson’s or Spearman's rank correlation coefficients. For principal component analysis, variables were natural log transformed and then zero-centered.

**Supplementary Tables:**

**Table S1:** List of mediators measured in the three data sets and used for hierarchical cluster analysis with the percentage of samples above and below the limit of. LLOQ, lower limit of quantification; ULOQ, upper limit of quantification.

| | GAinS (n=124) | | VANISH (n=155) | | LeoPARDS (n=484) | |
|---|---|---|---|---|---|---|
| Analyte | Percentage <LLOQ | Percentage >ULOQ | Percentage <LLOQ | Percentage >ULOQ | Percentage <LLOQ | Percentage >ULOQ |
| MCP-1 | 0.0 | 0.0 | 0.0 | 2.6 | 0.0 | 1.0 |
| IL-6 | 0.0 | 3.2 | 0.0 | 13.5 | 0.0 | 6.6 |
| IL-8 | 1.6 | 0.0 | 10.3 | 1.3 | 0.0 | 0.6 |
| IL-10 | 0.8 | 0.0 | 9.7 | 0.0 | 0.0 | 0.2 |
| IL-18 | 0.0 | 0.0 | 1.9 | 0.0 | 0.8 | 2.5 |
| CCL3 | 1.6 | 0.0 | 25.8 | 0.0 | - | - |
| IP-10 | 0.0 | 32.3 | 1.9 | 0.6 | - | - |
| IFN-γ | 0.0 | 1.6 | 55.5 | 0.6 | - | - |
| IL-1β | 3.2 | 0.0 | 67.1 | 0.0 | 8.9 | 0.0 |
| IL-2 | 26.6 | 0.0 | 66.5 | 0.0 | - | - |
| IL-17 | 0.0 | 0.0 | 66.5 | 0.0 | 1.9 | 0.0 |
| TNF-α | 0.0 | 0.0 | 78.1 | 0.0 | - | - |
| IFN-α | 36.3 | 0.0 | 71.6 | 0.0 | - | - |
| IL-12p70 | 3.2 | 0.0 | 88.4 | 0.0 | - | - |
| IL-4 | 0.0 | 0.0 | 87.7 | 0.0 | - | - |
| IL-1α | 13.7 | 0.8 | 77.4 | 0.0 | - | - |
| MCP-2 | 0.0 | 0.8 | - | - | - | - |
| IL-2R | 1.6 | 0.8 | - | - | - | - |
| SDF-1α | 0.0 | 3.2 | - | - | - | - |
| IL-27 | 0.0 | 0.0 | - | - | - | - |
| LIF | 5.6 | 0.0 | - | - | - | - |
| IL-5 | 6.5 | 0.0 | - | - | - | - |
| IL-7 | 0.0 | 0.0 | - | - | - | - |
| BLC | 0.0 | 0.0 | - | - | - | - |
| Eotaxin-2 | 4.0 | 0.0 | - | - | - | - |
| Eotaxin | 0.0 | 0.0 | - | - | - | - |
| IL-13 | 0.0 | 0.0 | - | - | - | - |
| IL-31 | 16.9 | 0.0 | - | - | - | - |
| SCF | 0.0 | 0.0 | - | - | - | - |
| G-CSF | 20.2 | 2.4 | - | - | - | - |
| GM-CSF | 0.0 | 0.0 | - | - | - | - |

| | | | | | | |
|---|---|---|---|---|---|---|
| HGF | 0.0 | 5.6 | - | - | - | - |
| MIP-1β | 0.8 | 0.0 | - | - | - | - |
| Eotaxin-3 | 0.0 | 0.0 | - | - | - | - |
| IL-9 | 0.0 | 0.0 | - | - | - | - |
| MIF | 0.0 | 0.8 | - | - | - | - |
| TNF-β | 4.0 | 0.0 | - | - | - | - |
| bNGF | 0.8 | 0.0 | - | - | - | - |
| MIP-3α | 0.0 | 0.0 | - | - | - | - |
| I-TAC | 0.0 | 0.0 | - | - | - | - |
| TRAIL | 0.0 | 0.0 | - | - | - | - |
| Fractalkine | 4.8 | 0.0 | - | - | - | - |
| GRO-α | 3.2 | 0.8 | - | - | - | - |
| IL-23 | 0.0 | 0.0 | - | - | - | - |
| MMP-1 | 4.0 | 0.0 | - | - | - | - |
| IL-15 | 31.5 | 0.0 | - | - | - | - |
| M-CSF | 0.0 | 12.1 | - | - | - | - |
| MCP-3 | 0.8 | 0.0 | - | - | - | - |
| MIG | 1.6 | 0.0 | - | - | - | - |
| IL-16 | 0.8 | 0.0 | - | - | - | - |
| IL-21 | 0.8 | 0.0 | - | - | - | - |
| IL-3 | 4.8 | 0.0 | - | - | - | - |
| CD40-ligand | 0.0 | 0.0 | - | - | - | - |
| FGF-2 | 3.2 | 0.0 | - | - | - | - |
| IL-22 | 2.4 | 0.8 | - | - | - | - |
| VEGF-A | 1.6 | 0.0 | - | - | - | - |
| TSLP | 0.0 | 0.0 | - | - | - | - |
| IL-20 | 0.0 | 0.0 | - | - | - | - |
| ENA-78 | 0.0 | 0.0 | - | - | - | - |
| CD30 | 0.0 | 0.0 | - | - | - | - |
| TNF-RII | 0.0 | 0.0 | - | - | - | - |
| BAFF | 7.3 | 0.0 | - | - | - | - |
| MDC | 0.0 | 0.0 | - | - | - | - |
| APRIL | 0.0 | 0.0 | - | - | - | - |
| Tweak | 0.0 | 0.0 | - | - | - | - |
| ICAM | - | - | 3.9 | 0.0 | 0.2 | 4.5 |
| CCL5 | - | - | 7.1 | 1.3 | - | - |
| sTNFR | - | - | 0.0 | 1.3 | 0.0 | 0.0 |
| MPO | - | - | 0.6 | 1.3 | 7.2 | 2.5 |
| Ang-2 | - | - | 1.3 | 0.0 | 1.4 | 1.7 |

**Table S2** Fit statistics for latent class models. Mean class probability is averaged over all class members. For LeoPARDS, we fit the full model with all class-predicting covariates and non-zero covariance between continuous indicators. For VANISH a reduced set of covariates was used to achieve convergence, and zero covariance between indicators was assumed based on model fit (see results for details). AIC= Akaike Information Criterion, BIC= Bayesian Information Criterion.

| | | | | | | AIC | BIC | Entropy |
|---|---|---|---|---|---|---|---|---|
| LeoPARDS | | Class 1 | Class 2 | Class 3 | Class 4 | | | |
| 1 class | # per class | 493 | | | | 25091 | 25404 | - |
| | Mean class probability | 1.00 | | | | | | |
| 2 classes | # per class | 283 | 210 | | | 23833 | 24339 | 0.87 |
| | Mean class probability | 0.97 | 0.96 | | | | | |
| 3 classes | # per class | 191 | 247 | 55 | | 23238 | 23936 | 0.91 |
| | Mean class probability | 0.94 | 0.96 | 0.98 | | | | |
| 4 classes | # per class | 151 | 132 | 156 | 54 | 22997 | 23887 | 0.88 |
| | Mean class probability | 0.95 | 0.93 | 0.90 | 0.98 | | | |
| VANISH | | | | | | | | |
| 1 class | # per class | 176 | | | | 7714 | 7874 | |
| | Mean class probability | 1.00 | | | | | | |
| 2 classes | # per class | 90 | 86 | | | 7378 | 7671 | 0.88 |
| | Mean class probability | 0.96 | 0.97 | | | | | |
| 3 classes | # per class | 46 | 67 | 63 | | 7298 | 7722 | 0.86 |
| | Mean class probability | 0.98 | 0.90 | 0.95 | | | | |
| 4 classes | # per class | 30 | 41 | 43 | 62 | 7281 | 7838 | 0.88 |
| | Mean class probability | 0.95 | 0.93 | 0.91 | 0.95 | | | |

**Table S3** Estimated class distribution, indicator means and separation for the LeoPARDS trial. In these models, all covariates were included and the residual variance of each indicator can differ across classes.

| | 2-class model | | | 3-class model | | | | 4-class model | | | | |
|---|---|---|---|---|---|---|---|---|---|---|---|---|
| | **Class 1** | **Class 2** | **Separation** | **Class 1** | **Class 2** | **Class 3** | **Separation** | **Class 1** | **Class 2** | **Class 3** | **Class 4** | **Separation** |
| Distribution (%) | 58 | 42 | | 39 | 50 | 11 | | 31 | 25 | 33 | 11 | |
| *Organ dysfunction* | | | | | | | | | | | | |
| $PaO_2/FiO_2$ ratio | 0.12 | -0.15 | 0.018 | 0.098 | -0.01 | -0.252 | 0.021 | 0.076 | 0.109 | -0.096 | -0.232 | 0.019 |
| Creatinine | -0.228 | 0.289 | 0.067 | -0.368 | 0.155 | 0.451 | 0.115 | -0.512 | 0.233 | 0.078 | 0.459 | 0.129 |
| Platelets | 0.238 | -0.319 | 0.078 | 0.304 | -0.087 | -0.58 | 0.131 | 0.378 | -0.335 | 0.119 | -0.627 | 0.152 |
| Bilirubin | -0.231 | 0.284 | 0.066 | -0.295 | 0.196 | 0.021 | 0.041 | -0.454 | 0.585 | -0.01 | 0.054 | 0.136 |
| *Inflammation markers* | | | | | | | | | | | | |
| IL-1β | -0.587 | 0.416 | 0.252 | -0.774 | -0.036 | 1.383 | 0.801 | -0.825 | -0.5 | 0.157 | 1.381 | 0.713 |
| IL-6 | -0.487 | 1.123 | 0.648 | -0.746 | 0.428 | 2.506 | 1.808 | -0.773 | -0.369 | 0.795 | 2.456 | 1.571 |
| IL-8 | -0.552 | 0.838 | 0.483 | -0.836 | 0.274 | 1.954 | 1.315 | -0.925 | -0.173 | 0.45 | 1.953 | 1.119 |
| IL-10 | -0.532 | 0.775 | 0.427 | -0.779 | 0.283 | 1.601 | 0.948 | -0.894 | 0.058 | 0.344 | 1.62 | 0.807 |
| IL-17 | -0.4 | 0.465 | 0.187 | -0.499 | 0.106 | 0.9 | 0.328 | -0.565 | 0.079 | 0.079 | 0.924 | 0.28 |
| IL-18 | -0.258 | 0.479 | 0.136 | -0.358 | 0.246 | 0.569 | 0.148 | -0.497 | 0.641 | -0.053 | 0.605 | 0.226 |
| *Leukocytes* | | | | | | | | | | | | |
| Myeloperoxidase | -0.373 | 0.303 | 0.114 | -0.531 | 0.125 | 0.461 | 0.17 | -0.64 | 0.194 | -0.008 | 0.49 | 0.172 |
| sICAM | -0.2 | 0.515 | 0.128 | -0.175 | 0.166 | 0.742 | 0.143 | -0.312 | 0.823 | -0.195 | 0.777 | 0.279 |
| *Endothelial injury* | | | | | | | | | | | | |
| Angiotensin-2 | -0.486 | 0.696 | 0.349 | -0.709 | 0.356 | 0.894 | 0.444 | -0.896 | 0.556 | 0.152 | 0.933 | 0.467 |
| *Cardiovascular* | | | | | | | | | | | | |
| Troponin | -0.017 | 0.018 | 0 | -0.034 | -0.017 | 0.157 | 0.007 | -0.09 | 0.126 | -0.128 | 0.18 | 0.018 |

| | | | | | | | | | | | | |
|---|---|---|---|---|---|---|---|---|---|---|---|---|
| B-natriuretic peptide | -0.368 | 0.07 | 0.048 | -0.346 | -0.184 | 0.361 | 0.091 | -0.496 | 0.166 | -0.359 | 0.359 | 0.126 |
| *Other markers* | | | | | | | | | | | | |
| sTNFr-1 | -0.414 | 0.56 | 0.237 | -0.659 | 0.333 | 0.705 | 0.331 | -0.865 | 0.404 | 0.229 | 0.719 | 0.355 |
| Lactate | -0.425 | 0.59 | 0.258 | -0.613 | 0.263 | 0.91 | 0.39 | -0.684 | 0.048 | 0.342 | 0.898 | 0.326 |
| MCP-1 (CCL2) | -0.533 | 0.873 | 0.494 | -0.753 | 0.262 | 1.944 | 1.237 | -0.817 | -0.087 | 0.369 | 1.966 | 1.041 |

**Table S4** Differences in class assignment for main and sensitivity analysis

| **Main analysis** | **Sensitivity analysis** | | |
|---|---|---|---|
| LeoPARDS | Class 1 | Class 2 | Class 3 |
| Class 1 | 180 | 11 | 0 |
| Class 2 | 5 | 228 | 14 |
| Class 3 | 0 | 0 | 55 |
| VANISH | Class 1 | Class 2 | |
| Class 1 | 87 | 3 | |
| Class 2 | 2 | 84 | |

**Table S5** Model coefficients from multinomial regression model using data from the LeoPARDS trial

| | Class 2 vs Class 1 | | Class 3 vs Class 1 | |
|---|---|---|---|---|
| | Log OR for a 1 SD increase | SE | Log OR for a 1 SD increase | SE |
| IL-6 | 1.404 | 0.356 | 3.803 | 0.771 |
| IL-8 | 2.477 | 0.472 | 4.09 | 0.735 |
| IL-10 | 1.942 | 0.346 | 2.514 | 0.639 |
| CCL2 | 1.314 | 0.328 | 3.976 | 0.853 |
| (baseline odds) | 2.236 | 0.301 | -8.264 | 1.952 |

**Table S6.** Comparison of inflammatory mediator concentrations between hierarchical clusters identified in the GAinS, VANISH and LeoPARDS datasets. The 16 mediators measured in both GAinS and VANISH are also shown in Table 3. Data are given as median and interquartile range in units of pg/mL and comparison has been made with the Mann-Whitney U test for two-group comparisons and the Kruskal-Wallis tests for three-group comparisons. FDR values in bold are those <0.05.

| | **GAinS** | | | **VANISH** | | | **LeoPARDS** | | | |
|---|---|---|---|---|---|---|---|---|---|---|
| Analyte | LC cluster (Low cytokine) | HC cluster (High cytokine) | FDR | LC cluster (Low cytokine) | HC cluster (High cytokine) | FDR | LC cluster (Low cytokine) | IC cluster (Intermediate cytokines) | HC cluster (High cytokine) | FDR |
| n | 70 | 54 | - | 71 | 84 | - | 191 | 208 | 85 | - |
| MCP-1 (CCL2) | 231.6 (124.2-402.6) | 444.1 (285.7-874.3) | **<1x10$^{-5}$** | 890.1 (533.9-1477.5) | 4410.2 (2061.5-9789.2) | **<1x10$^{-10}$** | 390.3 (279.5-623.9) | 867.2 (640.1-1333.3) | 3236.6 (1944-5282.3) | **<1x10$^{-50}$** |
| IL-6 | 980.3 (726.9-1516.6) | 1768.5 (1384.7-3005.1) | **<1x10$^{-5}$** | 337.4 (163.2-941.8) | 11535.1 (3394.2-34623.2) | **<1X10$^{-10}$** | 266.5 (100.4-575.6) | 1089.3 (434.3-3169.8) | 24761.5 (9608.8-40000) | **<1x10$^{-50}$** |
| IL-8 | 314.1 (187.7-411.5) | 624.4 (515-1052.1) | **<1x10$^{-10}$** | 32.1 (7.7-95.0) | 839.8 (186.3-3192.8) | **<1X10$^{-10}$** | 48.4 (30.4-91.6) | 226 (138.7-375.2) | 1931.8 (787-5332.2) | **<1x10$^{-50}$** |
| IL-10 | 156.9 (91.7-242.2) | 359.7 (239.3-701.9) | **<1x10$^{-10}$** | 10.2 (4.2-27.1) | 151.4 (44.7-397.5) | **<1x10$^{-10}$** | 25.4 (16.7-43) | 112.5 (70-196.9) | 432.2 (203.7-1049.9) | **<1x10$^{-50}$** |
| IL-18 | 607.5 (483.8-877.5) | 1173.5 (999.9-1551.3) | **<1x10$^{-10}$** | 354.3 (148.1-595.2) | 536.9 (335.7-823.4) | **<0.001** | 577.6 (381.3-905.3) | 912.5 (567.9-1363.3) | 896 (556.9-1642.5) | **<1x10$^{-5}$** |
| CCL3 | 72.1 (60-97.4) | 138.3 (108.6-176.9) | **<1x10$^{-10}$** | 11.9 (3.8-22.5) | 79.4 (32.0-144.0) | **<1x10$^{-10}$** | - | - | - | - |
| IP-10 | 332.6 (234.8-544.4) | 784 (409.5-787.5) | **<1x10$^{-05}$** | 443.3 (214.1-783.4) | 1248.6 (584.8-5498.9) | **<1x10$^{-5}$** | - | - | - | - |
| IFN-γ | 487.7 (385.9-609.2) | 977.5 (741.7-1520.9) | **<1x10$^{-10}$** | 3.6 (2.3-13.2) | 11.4 (3.6-175.0) | **<0.001** | - | - | - | - |
| IL-1β | 28 (12.7-65.3) | 151.7 (71.8-282.6) | **<1x10$^{-10}$** | 3.2 (2.8-3.8) | 3.8 (3.0-10.7) | **0.001** | 0.8 (0.5-1.3) | 1.4 (0.7-2.6) | 4.6 (1.9-10.3) | **<1x10$^{-20}$** |
| IL-2 | 24 (22-31.8) | 42.6 (32-59.5) | **<1x10$^{-05}$** | 4.5 (3.3-9.5) | 8.2 (4.5-14.5) | **0.02** | - | - | - | - |
| IL-17 | 488 (330.2-646.7) | 846.4 (699.9-1147.5) | **<1x10$^{-05}$** | 4.6 (2.7-7.7) | 6 (4.4-17.7) | **0.03** | 6.6 (4.6-9.7) | 9.6 (6.1-19) | 18 (7.7-48) | **<1x10$^{-10}$** |

| | | | | | | | | | | |
|---|---|---|---|---|---|---|---|---|---|---|
| TNF-α | 56.2 (34.5-82.4) | 138.2 (104.3-187) | **<1x10$^{-10}$** | 3.1 (2.4-5.3) | 3.9 (3.0-6.7) | **0.03** | - | - | - | - |
| IFN-α | 75 (75-104.6) | 172.9 (130.6-229.6) | **<1x10$^{-10}$** | 3.6 (2.8-4.6) | 3.9 (2.9-6.9) | 0.07 | - | - | - | - |
| IL-12p70 | 49.1 (24.7-75.1) | 151.7 (111.6-225) | **<1x10$^{-10}$** | 2.9 (2.4-4.4) | 3.7 (2.9-5.2) | 0.1 | - | - | - | - |
| IL-4 | 1856.2 (1402.9-2561.8) | 3319.6 (2659.1-4507.2) | **<1x10$^{-05}$** | 2.6 (2.5-5.4) | 4.9 (2.6-5.7) | 0.18 | - | - | - | - |
| IL-1α | 46.3 (27-63.3) | 153.8 (106.5-188.7) | **<1x10$^{-10}$** | 4.8 (2.8-15.9) | 5.3 (3.9-15.6) | 0.52 | - | - | - | - |
| MCP-2 | 76 (47.1-107.3) | 197.9 (129.8-291.5) | **<1x10$^{-10}$** | - | - | - | - | - | - | - |
| IL-2R | 12040.3 (7097.4-27527.9) | 29087.4 (18754.9-63393.4) | **<1x10$^{-05}$** | - | - | - | - | - | - | - |
| SDF-1α | 1761.3 (1212.4-2721.2) | 5347.9 (3486.1-7304.9) | **<1x10$^{-10}$** | - | - | - | - | - | - | - |
| IL-27 | 1192.6 (838.4-1707.3) | 2492.1 (1744.7-3450) | **<1x10$^{-10}$** | - | - | - | - | - | - | - |
| LIF | 11.2 (6.3-18.7) | 27.9 (19.3-46.8) | **<1x10$^{-10}$** | - | - | - | - | - | - | - |
| IL-5 | 58.5 (36.4-104.1) | 247 (175.7-343.4) | **<1x10$^{-10}$** | - | - | - | - | - | - | - |
| IL-7 | 19.5 (9.5-34.4) | 48.9 (34.6-78.7) | **<1x10$^{-5}$** | - | - | - | - | - | - | - |
| BLC | 255.5 (161.6-527.5) | 669.5 (373.8-1316.5) | **<1x10$^{-5}$** | - | - | - | - | - | - | - |
| Eotaxin-2 | 187.6 (126.4-278.1) | 371.3 (282.9-611.4) | **<1x10$^{-5}$** | - | - | - | - | - | - | - |
| Eotaxin | 96.4 (68.1-110.8) | 143.2 (114.4-186.8) | **<1x10$^{-10}$** | - | - | - | - | - | - | - |
| IL-13 | 95.9 | 178.5 | **<1x10$^{-10}$** | - | - | - | - | - | - | - |

| | | | | | | | | | | |
|---|---|---|---|---|---|---|---|---|---|---|
| | (69.6-135.3) | (154.4-251.1) | | | | | | | | |
| IL-31 | 585.5 (306.2-1033.1) | 1391.3 (889.9-2566.7) | **<1x10$^{-05}$** | - | - | - | - | - | - | - |
| SCF | 55.1 (37.4-74.3) | 137.3 (100.4-195) | **<1x10$^{-10}$** | - | - | - | - | - | - | - |
| G-CSF | 79.6 (39.5-160) | 323.5 (210.8-616.9) | **<1x10$^{-10}$** | - | - | - | - | - | - | - |
| GM-CSF | 162.7 (91.4-243.4) | 465.1 (309-621) | **<1x10$^{-10}$** | - | - | - | - | - | - | - |
| HGF | 1405.2 (1041.3-2311.3) | 2910.4 (1938.4-7442.3) | **<1x10$^{-05}$** | - | - | - | - | - | - | - |
| MIP-1β | 174.6 (123.7-258.1) | 375.5 (298.4-731.8) | **<1x10$^{-10}$** | - | - | - | - | - | - | - |
| Eotaxin-3 | 92.9 (67.9-120.7) | 176.3 (133.3-226.9) | **<1x10$^{-10}$** | - | - | - | - | - | - | - |
| IL-9 | 163.4 (117.3-229.8) | 411.7 (315.5-555) | **<1x10$^{-10}$** | - | - | - | - | - | - | - |
| MIF | 19.9 (13.1-29.6) | 39.9 (29.5-58.8) | **<1x10$^{-5}$** | - | - | - | - | - | - | - |
| TNF-β | 337.2 (207.6-560.3) | 986.9 (729.6-1537.1) | **<1x10$^{-10}$** | - | - | - | - | - | - | - |
| bNGF | 805.6 (655.3-963.6) | 1177.4 (1035.5-1468.6) | **<1x10$^{-5}$** | - | - | - | - | - | - | - |
| MIP-3α | 801.8 (475.8-1308) | 1670.3 (1235.2-2902.5) | **<1x10$^{-5}$** | - | - | - | - | - | - | - |
| I-TAC | 319.5 (220.5-488.3) | 845.9 (633.2-1306.5) | **<1x10$^{-10}$** | - | - | - | - | - | - | - |
| TRAIL | 128.4 (90.9-167.3) | 298.1 (228-404.5) | **<1x10$^{-10}$** | - | - | - | - | - | - | - |
| Fractalkine | 13.5 (6.6-17.1) | 37.5 (28.7-56.7) | **<1x10$^{-10}$** | - | - | - | - | - | - | - |
| GRO-α | 71.6 (29.2-101) | 187.6 (143.1-255.9) | **<1x10$^{-10}$** | - | - | - | - | - | - | - |

| | | | | | | | | | | |
|---|---|---|---|---|---|---|---|---|---|---|
| IL-23 | 1712.9 (1172.1-2420.7) | 3119.8 (2392-3993.7) | **$<1x10^{-10}$** | - | - | - | - | - | - | - |
| MMP-1 | 51.5 (28.4-86.1) | 135 (101.9-215.8) | **$<1x10^{-10}$** | - | - | - | - | - | - | - |
| IL-15 | 13.4 (13.4-29.2) | 64.8 (42.2-108.3) | **$<1x10^{-10}$** | - | - | - | - | - | - | - |
| M-CSF | 1232.2 (767.4-1662.8) | 3026.1 (2198.1-4189.1) | **$<1x10^{-10}$** | - | - | - | - | - | - | - |
| MCP-3 | 184.3 (124.1-251.8) | 446.7 (329-618) | **$<1x10^{-10}$** | - | - | - | - | - | - | - |
| MIG | 99.4 (64.2-170.2) | 401.1 (245.3-769.5) | **$<1x10^{-10}$** | - | - | - | - | - | - | - |
| IL-16 | 584.4 (363.6-820.8) | 990.7 (670.7-1328.2) | **$<1x10^{-5}$** | - | - | - | - | - | - | - |
| IL-21 | 149.5 (72.9-213.1) | 362.9 (277.8-518.3) | **$<1x10^{-10}$** | - | - | - | - | - | - | - |
| IL-3 | 93.6 (64-138.4) | 398.7 (254.7-628.3) | **$<1x10^{-10}$** | - | - | - | - | - | - | - |
| CD40-ligand | 170.4 (128-266.5) | 467.2 (315.5-644) | **$<1x10^{-10}$** | - | - | - | - | - | - | - |
| FGF-2 | 293 (214.6-430.1) | 665.9 (547.9-808.1) | **$<1x10^{-10}$** | - | - | - | - | - | - | - |
| IL-22 | 389.8 (262.9-517.9) | 1121 (801.5-1551.4) | **$<1x10^{-10}$** | - | - | - | - | - | - | - |
| VEGF-A | 326.3 (237.9-501.1) | 888.3 (696.3-1294.5) | **$<1x10^{-10}$** | - | - | - | - | - | - | - |
| TSLP | 132.7 (97.7-185.5) | 268.1 (220.9-416.9) | **$<1x10^{-10}$** | - | - | - | - | - | - | - |
| IL-20 | 202.5 (134-296.7) | 435.4 (370.5-566.5) | **$<1x10^{-10}$** | - | - | - | - | - | - | - |
| ENA-78 | 202.2 (127.2-280.4) | 353.3 (238.3-484.3) | **$<1x10^{-5}$** | - | - | - | - | - | - | - |
| CD30 | 1439.1 | 3211.7 | **$<1x10^{-5}$** | - | - | - | - | - | - | - |

| | | | | | | | | | | |
|---|---|---|---|---|---|---|---|---|---|---|
| | (945.3-2274.9) | (2337.3-4428.7) | | | | | | | | |
| TNF-RII | 172.4 (118.1-216.2) | 284.8 (233.7-405.4) | **<1x10$^{-10}$** | - | - | - | - | - | - | - |
| BAFF | 139.7 (81.3-177.8) | 362.4 (278.5-593.4) | **<1x10$^{-10}$** | - | - | - | - | - | - | - |
| MDC | 426 (281.1-526.7) | 677.9 (577.1-835.8) | **<1x10$^{-10}$** | - | - | - | - | - | - | - |
| APRIL | 7304.7 (3887.3-10704) | 18394.9 (11938.9-33962.1) | **<1x10$^{-10}$** | - | - | - | - | - | - | - |
| Tweak | 4557.8 (3428.9-6340.9) | 9792.2 (7997.2-12516.4) | **<1x10$^{-10}$** | - | - | - | - | - | - | - |
| sTNFr1 | - | - | - | 4087.6 (2085.2-6517.3) | 7951.3 (4984.3-11821.7) | **<1x10$^{-5}$** | 5957.4 (3922.7-9991) | 13815.3 (9175-20641.9) | 14351.2 (9708.5-23711.7) | **<1x10$^{-20}$** |
| MPO | - | - | - | 292057.8 (152041.4-461508.9) | 633615.7 (318953.6-1211865.2) | **4.9x10$^{-5}$** | 269062 (175736.1-483624.4) | 532337.2 (322949.8-1206784.4) | 463795 (234525.1-1349336.9) | **<1x10$^{-10}$** |
| Ang-2 | - | - | - | 3172.3 (1493.3-5478.9) | 6384.5 (3699.9-11171.1) | **1.6x10$^{-5}$** | 3217.1 (1910.4-5127.1) | 8315.1 (4536.5-16484.4) | 9777.4 (5716-22919.1) | **<1x10$^{-20}$** |
| ICAM | - | - | - | 232608.7 (160025.8-370906.2) | 321250.4 (209874.2-519756.3) | **0.002** | 282668.1 (184974.8-440678.7) | 328433.4 (186577.9-582882.8) | 402553.3 (242763.6-844714.2) | **<0.001** |
| CCL5 | - | - | - | 5969.7 (1702.9-15472.4) | 4621.4 (1151.1-15554.8) | 0.5 | - | - | - | - |

**Table S7:** Comparison of the LCA probabilities of class membership between those patients similarly classified by LCA and HCA and those that were not. Probabilities are given as medians and inter-quartile range and p-values were obtained with the Mann-Whitney U test. P-values in bold are those <0.05.

| LCA group | LCA probabilities VANISH | | | LCA probabilities LeoPARDS | | |
|---|---|---|---|---|---|---|
| | HCA match | HCA mismatch | p-value | HCA match | HCA mismatch | p-value |
| 1 | 1.000 (0.996-1.000) | 0.957 (0.681-0.999) | **0.001** | 1.000 (0.983-1.000) | 0.856 (0.642-0.990) | **<0.001** |
| 2 | 1.000 (0.993-1.000) | 0.980 (0.980-0.997) | 0.08 | 0.999 (0.988-1.000) | 0.983 (0.871-0.999) | **<0.001** |
| 3 | - | - | - | 1.000 (0.998-1.000) | 0.994 (0.994-0.994) | 0.28 |

**Table S8.** Comparison of baseline variables and clinical outcomes between clusters from the hierarchical cluster analysis models in GAinS, VANISH and LeoPARDS. A subset of the tables is also shown as Table 4. Continuous variables have been compared with the Mann-Whitney U test or Kruskal-Wallis test and categorical variables with the chi-squared test or Fisher's exact test (if number of events <10). P-values in bold are those <0.05. No adjustment has been made for multiple comparisons. (APACHE=Acute Physiology and Chronic Health Evaluation, COPD=Chronic Obstructive Pulmonary Disease, NYHA= New York Heart Association, GCS=Glasgow Coma Scale, IV=intravenous, IQR=interquartile range, ICU=Intensive Care Unit). * Organ failure is defined as having a Sequential Organ Failure Assessment (SOFA) score of 3 or more. † In VANISH acute renal failure was defined as having acute kidney injury stage 3.

| | **GAinS** | | | **VANISH** | | | **LeoPARDS** | | | |
|---|---|---|---|---|---|---|---|---|---|---|
| | LC cluster (Low cytokine) | HC cluster (High cytokine) | p-value | LC cluster (Low cytokine) | HC cluster (High cytokine) | p-value | LC cluster (Low cytokine) | IC cluster (Intermediate cytokine) | HC cluster (High cytokine) | p-value |
| n | 70 | 54 | - | 71 | 84 | - | 191 | 208 | 85 | - |
| Age median (IQR), y | 68 (52-77) | 69 (62-76) | 0.5 | 65 (54-77) | 64 (52-77) | 0.46 | 68 (58-77) | 69 (60.3-76) | 66 (52-74) | 0.13 |
| Men, No./total (%) | 42/70 (60) | 38/54 (70) | 0.23 | 49/71 (69) | 52/84 (62) | 0.36 | 106/191 (55) | 116/208 (56) | 46/85 (54) | 0.97 |
| Caucasian ethnicity, No./total (%) | 67/70 (96) | 51/52 (98) | 0.64 | 60/71 (85) | 68/84 (81) | 0.56 | 178/191 (93) | 193/208 (93) | 81/85 (95) | 0.73 |
| Recent surgical history, No./total (%) | 7/69 (10) | 11/54 (20) | 0.13 | 14/71 (20) | 11/84 (13) | 0.26 | 53/191 (28) | 86/208 (41) | 37/85 (44) | **0.01** |
| APACHE II score, median (IQR) | 15 (11-20) | 16 (12-21) | 0.23 | 23 (18-28) | 24 (20-31) | 0.07 | 24 (21-30) | 26 (22-31) | 26 (21.5-32.5) | **0.02** |
| Baseline total SOFA score, median (IQR) | 6 (3-7) | 6 (4-9) | 0.11 | - | - | - | 10 (7-11) | 10 (8-13) | 11 (8-14) | **<0.001** |
| *Pre-existing conditions, No./total (%)* | | | | | | | | | | |
| Ischemic heart disease | 11/70 (16) | 7/54 (13) | 0.8 | 17/71 (24) | 8/84 (10) | **0.02** | 29/191 (15) | 26/208 (13) | 16/85 (19) | 0.37 |
| Severe COPD | - | - | - | 5/71 (7) | 3/84 (4) | 0.47 | 13/191 (7) | 6/208 (3) | 4/85 (5) | 0.18 |
| COPD | 22/70 (31) | 12/54 (22) | 0.25 | - | - | - | - | - | - | - |
| Chronic kidney failure | 12/70 (17) | 5/54 (9) | 0.29 | 2/71 (3) | 5/84 (6) | 0.45 | 17/191 (9) | 15/208 (7) | 3/85 (4) | 0.28 |
| Cirrhosis | 1/70 (1) | 1/54 (2) | 1 | 0/71 (0) | 10/84 (12) | **0.002** | 5/191 (3) | 4/208 (2) | 0/85 (0) | 0.37 |

| | | | | | | | | | | |
|---|---|---|---|---|---|---|---|---|---|---|
| Cancer | 9/70 (13) | 8/54 (15) | 0.8 | 6/71 (9) | 12/84 (14) | 0.32 | - | - | - | - |
| Immunocompromised | - | - | - | 0/71 (0) | 10/84 (12) | **0.002** | 14/191 (7) | 19/208 (9) | 11/85 (13) | 0.33 |
| Diabetes | 10/70 (14) | 7/54 (13) | 1 | 14/71 (20) | 17/84 (20) | 0.94 | 41/191 (21) | 49/208 (24) | 14/85 (16) | 0.41 |
| Cardiac Failure | 2/70 (3) | 0/54 (0) | 0.5 | - | - | - | 16/191 (8) | 20/208 (10) | 8/85 (9) | 0.91 |
| NYHA Heart failure class IV | - | - | - | - | - | - | 1/191 (1) | 4/208 (2) | 0/85 (0) | 0.34 |
| *Organ failure, No./total (%)** | | | | | | | | | | |
| Cardiovascular | 25/70 (36) | 25/54 (46) | 0.23 | - | - | - | 190/191 (99) | 207/208 (100) | 84/85 (99) | 0.77 |
| Respiratory | 10/70 (14) | 12/54 (22) | 0.25 | 21/70 (30) | 35/83 (42) | 0.12 | 78/189 (41) | 69/208 (33) | 40/85 (47) | 0.06 |
| Kidney † | 12/70 (17) | 11/54 (20) | 0.65 | 13/71 (18) | 22/84 (26) | 0.24 | 13/191 (7) | 24/207 (12) | 14/84 (17) | **0.04** |
| Liver | 0/70 (0) | 2/54 (4) | 0.19 | 4/60 (7) | 6/77 (8) | 1 | 3/187 (2) | 8/202 (4) | 2/85 (2) | 0.37 |
| Hematological | 0/70 (0) | 1/54 (2) | 0.44 | 2/69 (3) | 7/82 (9) | 0.18 | 4/190 (2) | 12/207 (6) | 10/84 (12) | **0.004** |
| Neurological | 3/70 (4) | 1/54 (2) | 0.63 | 24/69 (35) | 26/80 (33) | 0.77 | 91/167 (54) | 85/176 (48) | 38/67 (57) | 0.37 |
| *Physiological variables, median (IQR)* | | | | | | | | | | |
| Mean arterial pressure, mmHg | 64 (55-71) | 60 (56-70) | 0.67 | 69 (61-78) | 69 (62-75) | 0.41 | 75 (70-81) | 73.5 (67-79) | 71 (65-75.5) | **0.002** |
| Highest heart rate, beats/min | 110 (97-130) | 116 (100-134) | 0.35 | - | - | **-** | | | | |
| Lowest heart rate, beats/min | 76 (65-86) | 82 (74-88) | **0.008** | 85 (75-101) | 100 (90-118) | **<0.001** | 86 (73-100) | 98 (84-113) | 110 (92-121) | **<1x10$^{-13}$** |
| Central venous pressure, mmHg | - | - | - | 13 (7-19) | 14 (10-19) | 0.35 | 10 (8-15) | 12 (9-16) | 12 (9-14.3) | 0.33 |
| Lactate, mmol/L | 2.2 (1.6-3.4) | 2.3 (1.4-3.5) | 0.98 | 1.8 (1.2-2.8) | 3.0 (2.0-5.0) | **<0.001** | 1.5 (1-2.2) | 2.7 (1.8-3.9) | 3.9 (2.5-6.2) | **<1x10$^{-28}$** |
| $PaO_2/FiO_2$, mmHg | 154 (100-226) | 120 (80-186) | **0.048** | 271 (160-332) | 160 (109-260) | **<0.001** | 212 (159-297) | 226 (160-301.9) | 194 (123-264) | **0.01** |
| Creatinine, mg/dL | 1.1 (0.7-1.6) | 1.1 (0.8-2.0) | 0.39 | 1.1 (0.8-1.8) | 1.5 (1.0-2.8) | **0.004** | 1.2 (0.8-1.9) | 1.8 (1.2-2.6) | 1.8 (1.4-2.9) | **<1x10$^{-10}$** |
| Bilirubin, mg/dL | 0.6 (0.4-1.0) | 0.6 (0.5-1.4) | 0.23 | 0.6 (0.4-1.2) | 1.0 (0.6-2.3) | **0.005** | 0.7 (0.4-1.1) | 1 (0.5-1.9) | 1 (0.5-1.8) | **<1x10$^{-6}$** |
| Platelets, ×$10^3$/μL | 194 (153-273) | 190 (154-257) | 0.8 | 206 (151-342) | 170 (96-264) | **0.03** | 244 (181-351) | 208 (131-299) | 164 (87-241) | **<1x10$^{-7}$** |
| GCS | 15 (15-15) | 15 (15-15) | 0.45 | 13 (5-15) | 14 (4-15) | 0.64 | 6 (3-15) | 10.5 (3-15) | 3 (3-15) | 0.31 |
| Bicarbonate, mmol/L | 24.0 (20.0-28.6) | 22.3 (20.0-24.0) | **0.034** | - | - | - | - | - | - | - |

| | | | | | | | | | | |
|---|---|---|---|---|---|---|---|---|---|---|
| Highest white cell count, x$10^3$ / µL | 13.0 (9.9-17.7) | 13.8 (9.0-18.4) | 0.8 | - | - | - | - | - | - | - |
| Highest temperature, °C | 37.3 (36.9-37.8) | 37.9 (37.1-38.5) | **0.009** | - | - | - | - | - | - | - |
| Mechanical ventilation, No./total (%) | 47/70 (67) | 34/54 (63) | 0.63 | 39/71 (55) | 43/84 (51) | 0.64 | 151/191 (79) | 166/208 (80) | 73/85 (86) | 0.39 |
| Renal replacement therapy, No./total (%) | 5/70 (7) | 6/54 (11) | 0.53 | 2/71 (3) | 3/84 (4) | 1 | 15/191 (8) | 47/208 (23) | 24/85 (28) | **<0.001** |
| Volume of IV fluid in previous 4 h, median (IQR), mL | - | - | - | 850 (439-1499) | 1250 (610-2050) | **0.03** | 673 (411-1000) | 725 (366.3-1111.3) | 1087 (599-1568) | **$<1x10^{-4}$** |
| Norepinephrine dose at randomization, median (IQR), µg/kg/min | - | - | - | 0.13 (0.08-0.20) | 0.17 (0.10-0.31) | **0.03** | 0.2 (0.1-0.4) | 0.3 (0.2-0.5) | 0.5 (0.3-0.8) | **$<1x10^{-11}$** |
| *Source of infection, No./total (%)* | | | | | | | | | | |
| Lung | 53/70 (76) | 39/54 (72) | 0.66 | 31/69 (45) | 34/83 (41) | 0.62 | 105/190 (55) | 62/208 (30) | 22/85 (26) | **<0.001** |
| Abdomen | 17/70 (24) | 15/54 (28) | | 12/69 (17) | 21/83 (25) | 0.24 | 48/190 (25) | 89/208 (43) | 40/85 (47) | **<0.001** |
| Soft tissue or line | - | - | | 2/69 (3) | 3/83 (4) | 1 | 11/190 (6) | 7/208 (3) | 7/85 (8) | 0.19 |
| Other | - | - | | 24/69 (35) | 25/83 (30) | 0.54 | 26/190 (14) | 50/208 (24) | 16/85 (19) | **0.03** |
| Vasopressor or inotrope administration, No./total (%) | 25/70 (36) | 26/54 (48) | 0.23 | - | - | - | - | - | - | - |
| Inotropes No./total (%) | - | - | - | 2/71 (3) | 20/84 (24) | **<0.001** | - | - | - | - |
| SRS1 No./total (%) | 35/66 (53) | 29/49 (59) | 0.51 | 18/68 (26) | 51/81 (63) | **<0.001** | - | - | - | - |
| SRS2 No./total (%) | 31/66 (47) | 20/49 (41) | | 50/68 (74) | 30/81 (37) | | - | - | - | - |
| **Outcomes** | | | | | | | | | | |
| 28-d Mortality, No./total (%) | 13/69 (19) | 14/54 (26) | 0.35 | 14/71 (20) | 30/84 (36) | **0.03** | 46/190 (24) | 71/208 (34) | 38/85 (45) | **0.002** |
| ICU mortality, No./total (%) | 10/69 (14) | 14/54 (26) | 0.11 | 12/71 (17) | 24/84 (29) | 0.09 | 48/191 (25) | 61/208 (29) | 38/85 (45) | **0.004** |
| Hospital mortality, No./total (%) | 19/69 (27) | 17/54 (26) | 0.63 | 14/71 (20) | 28/84 (33) | 0.06 | 54/190 (28) | 75/208 (36) | 39/85 (46) | **0.02** |
| 3-month Mortality, No./total (%) | 21/69 (30) | 17/54 (31) | 0.9 | - | - | - | 57/188 (30) | 78/208 (38) | 42/85 (49) | **0.01** |

| | | | | | | | | | | |
|---|---|---|---|---|---|---|---|---|---|---|
| 6-month Mortality, No./total (%) | 23/69 (33) | 19/54 (35) | 0.82 | - | - | - | 60/188 (32) | 86/208 (41) | 43/85 (51) | **0.01** |
| Kidney failure, No./total (%) | 16/70 (23) | 14/54 (26) | 0.69 | 28/71 (39) | 43/84 (51) | 0.14 | - | - | - | - |
| Kidney failure free days, median (IQR), days | - | - | - | 22 (5-26) | 8 (0-22) | **0.02** | - | - | - | - |
| Duration of renal replacement therapy, median (IQR), days | - | - | - | - | - | - | 0 (0-0) | 0 (0-4) | 1 (0-5) | **$<1x10^{-8}$** |
| No. weaned from vasopressors for >24 h, No./total (%) | - | - | - | 68/71 (96) | 73/84 (87) | 0.06 | - | - | - | - |
| Time to shock reversal, median (IQR), hours | - | - | **-** | 43 (24-84) | 36 (16-73) | 0.24 | - | - | - | - |
| Duration of inotrope/vasopressor support, median(IQR), days | 0 (0-3) | 1.5 (0-5) | **0.026** | - | - | - | - | - | - | - |
| Catecholamine free days, median (IQR), days | - | - | - | - | - | - | 24 (4.5-26) | 23 (0-26) | 14 (0-25) | **<0.001** |
| Duration of mechanical ventilation, median (IQR), days | 3 (1-8) | 4 (1-12) | 0.2 | 6 (3-11) | 5 (2-11) | 0.27 | - | - | - | - |
| Ventilator free days, median (IQR), days | - | - | - | - | - | - | 20 (0.5-27) | 19 (0-26) | 2 (0-20) | **$<1x10^{-5}$** |
| Mean total SOFA score over ICU stay, median (IQR) | - | - | - | 3.9 (3.0-5.5) | 5.7 (4.0-9.4) | **<0.001** | 4.1 (2.9-6.1) | 5.9 (3.9-9) | 7.3 (4.3-13) | **$<1x10^{-12}$** |
| ICU length of stay, median (IQR), days | 6 (4-11) | 8 (4-14) | 0.3 | 6 (3-12) | 6 (3-12) | 0.72 | 7.9 (4-14.6) | 8 (3.7-13.6) | 7.9 (2.4-14.8) | 0.48 |
| Hospital length of stay, median (IQR), days | 19.0 (9.0-31.0) | 16.5 (10.0-32.0) | 0.95 | 21 (10-43) | 15 (6-33) | **0.03** | 22.6 (11.8-40.3) | 22.4 (12.2-43.4) | 17.7 (4.7-51.3) | 0.28 |

**Table S9:** Differential cytokine abundance analysis between the SRS1 and SRS2 transcriptomic sub-phenotypes in GAinS and VANISH. Concentrations in each group are given as median and interquartile range in units of pg/mL. Comparisons are made with the Mann-Whitney U test. FDR values in bold are those <0.05. A fold change larger than one indicates higher median level in SRS1.

| Mediator | GAinS | | | | VANISH | | | |
|---|---|---|---|---|---|---|---|---|
| | SRS1 | SRS2 | Fold Change | FDR | SRS1 | SRS2 | Fold Change | FDR |
| n | 64 | 51 | - | **-** | 69 | 80 | - | - |
| MCP-1 | 400.2 (218.8-739.1) | 223.2 (118.8-369.3) | 1.79 | **0.005** | 2998.9 (1283.9-7679.1) | 1289.8 (595.2-3636.6) | 2.33 | **<0.001** |
| IL-6 | 1768.5 (1185-2843.8) | 1043 (716.3-1436.7) | 1.7 | **0.0008** | 8253.6 (1557.1-32205.6) | 497.1 (177.1-2931) | 16.6 | **$<1x10^{-7}$** |
| IL-8 | 516.4 (376.4-633.1) | 386.7 (195.4-556.8) | 1.34 | **0.03** | 288.2 (54.8-2446.2) | 93.5 (26-588.3) | 3.08 | **0.01** |
| IL-10 | 245.9 (147.5-418.9) | 195.9 (99.6-354.1) | 1.25 | 0.34 | 107.2 (27.7-375.4) | 17.8 (6-57.1) | 6.01 | **$<1x10^{-5}$** |
| IL-18 | 875.9 (599.1-1279.7) | 872.6 (557.1-1136.2) | 1 | 0.62 | 462.1 (236.9-665.8) | 471.4 (217.7-881.8) | 0.98 | 0.52 |
| CCL3 | 106.8 (80.1-156.3) | 83 (61.9-121.6) | 1.29 | **0.03** | 54.1 (15.4-135.3) | 20.2 (8.6-53.5) | 2.67 | **<0.01** |
| IP-10 | 544.6 (341.1-787.5) | 359.5 (243.5-477.7) | 1.52 | **0.02** | 834 (344-3496.7) | 581.5 (264.6-1322.3) | 1.43 | 0.14 |
| IFN-γ | 668 (504.9-976.3) | 572.3 (423.9-908.5) | 1.17 | 0.31 | 4.9 (2.4-45.2) | 6.3 (2.4-29.8) | 0.78 | 0.81 |
| IL-1β | 61.4 (28.3-143.2) | 58.8 (19-136.8) | 1.05 | 0.48 | 3.6 (2.8-5.9) | 3.6 (2.8-5.5) | 1 | 0.84 |
| IL-2 | 33.4 (23.7-42.8) | 25.6 (22-42.3) | 1.3 | 0.27 | 5.1 (3.3-12.2) | 6.4 (4.1-13.6) | 0.8 | 0.78 |
| IL-17 | 713.5 (546.4-1133.8) | 602.4 (366.6-943.9) | 1.18 | 0.1 | 4.6 (3.1-12.8) | 6.3 (4.6-15.3) | 0.73 | 0.15 |
| TNF-α | 84.1 (54.5-131) | 82.9 (49.7-138.2) | 1.01 | 0.62 | 3.1 (2.4-5.8) | 4 (3-6.1) | 0.78 | 0.41 |
| IFN-α | 123 (75-155.7) | 89.9 (75-167) | 1.37 | 0.4 | 3.7 (2.7-6.4) | 3.7 (3-5.4) | 1 | 0.81 |
| IL-12p70 | 84.8 (46.1-144.2) | 96.1 (35.3-169.3) | 0.88 | 0.74 | 2.9 (2.4-4.7) | 3.7 (2.9-4.5) | 0.79 | 0.52 |
| IL-4 | 2540.6 (1683-3159.6) | 2569.2 (1551.7-3640.8) | 0.99 | 0.85 | 4.9 (2.6-5.7) | 4.9 (2.2-5.7) | 1 | 0.44 |
| IL-1α | 69 (46.6-154.6) | 65.8 (29.9-138.5) | 1.05 | 0.46 | 4.8 (3.1-15.6) | 5.3 (3.4-15.6) | 0.9 | 0.94 |
| MCP-2 | 123 | 103.9 | 1.18 | 0.26 | - | - | - | - |

| | | | | | | | | |
|---|---|---|---|---|---|---|---|---|
| | (78.8-200.9) | (56.9-173.8) | | | | | | |
| IL-2R | 28647.8 (12275.6-42128.9) | 15755 (7777.3-24241.3) | 1.82 | **0.02** | - | - | - | - |
| SDF-1α | 2720 (1664.9-4602.5) | 3066 (1482.9-5347.9) | 0.89 | 1 | - | - | - | - |
| IL-27 | 1581.3 (1087.1-2482.5) | 1658 (957.1-2562.5) | 0.95 | 0.74 | - | - | - | - |
| LIF | 20.9 (11.7-31.4) | 15.8 (6.1-27.9) | 1.32 | 0.1 | - | - | - | - |
| IL-5 | 123.2 (62.7-250.2) | 103.5 (46.3-197.1) | 1.19 | 0.34 | - | - | - | - |
| IL-7 | 36.4 (21.3-54.8) | 30.3 (11.8-50.3) | 1.2 | 0.25 | - | - | - | - |
| BLC | 422.8 (236.1-1294.1) | 324.2 (182.2-589.1) | 1.3 | 0.1 | - | - | - | - |
| Eotaxin-2 | 262.1 (160.1-389.3) | 274.1 (157.3-462.3) | 0.96 | 0.74 | - | - | - | - |
| Eotaxin | 115.2 (90-153) | 103.5 (74.2-120.8) | 1.11 | 0.13 | - | - | - | - |
| IL-13 | 142.2 (107.1-182) | 114.3 (71.8-201.1) | 1.25 | 0.39 | - | - | - | - |
| IL-31 | 1056.3 (668.1-1818.2) | 657.9 (318.4-1214.2) | 1.61 | **0.04** | - | - | - | - |
| SCF | 88.3 (58.7-123.1) | 67.1 (43.2-119.3) | 1.32 | 0.26 | - | - | - | - |
| G-CSF | 202.3 (75.7-398.3) | 113.3 (39.5-264.8) | 1.79 | 0.13 | - | - | - | - |
| GM-CSF | 243.4 (163.4-471.3) | 286.3 (140.3-521.8) | 0.85 | 0.74 | - | - | - | - |
| HGF | 2521.9 (1512.6-5254.2) | 1584.4 (1089.4-2279) | 1.59 | **0.007** | - | - | - | - |
| MIP-1β | 285.6 (184.5-452.2) | 245.9 (130.1-342.9) | 1.16 | 0.2 | - | - | - | - |
| Eotaxin-3 | 129 (84.1-167.3) | 113.3 (84.8-170) | 1.14 | 0.49 | - | - | - | - |
| IL-9 | 285.6 (193.9-367.1) | 212.9 (123.4-369.7) | 1.34 | 0.2 | - | - | - | - |
| MIF | 29.6 (18.9-44.7) | 22.1 (14.8-39.3) | 1.34 | 0.37 | - | - | - | - |
| TNF-β | 639.1 (395.9-920.8) | 599.7 (243.1-929) | 1.07 | 0.41 | - | - | - | - |
| bNGF | 1018.7 (808.8-1174.3) | 909.1 (680.7-1243.4) | 1.12 | 0.62 | - | - | - | - |
| MIP-3α | 1280.7 (940.6-2243.3) | 1052.4 (510.3-1482.6) | 1.22 | **0.03** | - | - | - | - |
| I-TAC | 521.6 (336.5-858.4) | 496.5 (235.8-821.2) | 1.05 | 0.32 | - | - | - | - |
| TRAIL | 187.3 (122.8-283.1) | 184.9 (112.5-261.9) | 1.01 | 0.74 | - | - | - | - |
| Fractalkine | 18.2 | 17.3 | 1.05 | 0.4 | - | - | - | - |

| | | | | | | | | |
|---|---|---|---|---|---|---|---|---|
| | (12.7-40.7) | (8.9-30.2) | | | | | | |
| GRO-α | 131.8 (77.1-187.3) | 91.5 (29.9-128.9) | 1.44 | 0.1 | - | - | - | - |
| IL-23 | 2345.9 (1536-3165.4) | 2395.8 (1390.6-3269.9) | 0.98 | 1 | - | - | - | - |
| MMP-1 | 107.8 (67.8-174.6) | 56.6 (29.3-140.8) | 1.91 | **0.03** | - | - | - | - |
| IL-15 | 32 (13.4-63.8) | 19.6 (13.4-62.5) | 1.64 | 0.49 | - | - | - | - |
| M-CSF | 1705.7 (1215.7-2730.3) | 1669.5 (941.6-2669.6) | 1.02 | 0.66 | - | - | - | - |
| MCP-3 | 285.7 (184.1-494.7) | 245.4 (154.5-388.9) | 1.16 | 0.32 | - | - | - | - |
| MIG | 194.2 (122.4-460) | 150.1 (72.4-327.4) | 1.29 | 0.16 | - | - | - | - |
| IL-16 | 758.9 (495.8-1049.5) | 672.5 (394.6-1057.2) | 1.13 | 0.66 | - | - | - | - |
| IL-21 | 245.8 (158.1-340.3) | 203.5 (80.3-297.4) | 1.21 | 0.25 | - | - | - | - |
| IL-3 | 147.5 (94.4-373.8) | 139.2 (73.2-344.1) | 1.06 | 0.46 | - | - | - | - |
| CD40-ligand | 299.5 (167.5-531.6) | 270.9 (147.3-433) | 1.11 | 0.45 | - | - | - | - |
| FGF-2 | 486.8 (299.1-677.4) | 430.1 (259.5-586.2) | 1.13 | 0.4 | - | - | - | - |
| IL-22 | 583 (419.3-1132.6) | 498.9 (286.1-1052.4) | 1.17 | 0.39 | - | - | - | - |
| VEGF-A | 517.3 (383.8-825.5) | 505.7 (245.5-826.8) | 1.02 | 0.48 | - | - | - | - |
| TSLP | 187.4 (134.9-296.6) | 187.4 (104.9-250.4) | 1 | 0.39 | - | - | - | - |
| IL-20 | 293.4 (184-474.3) | 313.6 (187.1-404.9) | 0.94 | 0.74 | - | - | - | - |
| ENA-78 | 285.5 (202.7-447.5) | 225.6 (155.4-337) | 1.27 | 0.13 | - | - | - | - |
| CD30 | 2200.4 (1388.8-4040.6) | 2236.4 (1248.3-3447) | 0.98 | 0.9 | - | - | - | - |
| TNF-RII | 220.9 (168.9-295.5) | 207.2 (129.3-269.9) | 1.07 | 0.32 | - | - | - | - |
| BAFF | 226.1 (146.3-384.5) | 193.5 (105.3-352.2) | 1.17 | 0.46 | - | - | - | - |
| MDC | 508.7 (367.4-658.5) | 534.7 (355.8-728.8) | 0.95 | 0.97 | - | - | - | - |
| APRIL | 10810 (7397.2-23132.6) | 9838 (4080.1-14628.1) | 1.1 | 0.16 | - | - | - | - |
| Tweak | 6342 (4444-9932.6) | 6067 (4162.9-9199.5) | 1.05 | 0.48 | - | - | - | - |
| ICAM | - | - | - | - | 286696.2 (209540.6-480708.6) | 257379.4 (168244-420958.9) | 1.11 | 0.48 |
| CCL5 | - | - | - | - | 4650.5 | 5030.1 | 0.92 | 0.96 |

| | | | | | | | | |
|---|---|---|---|---|---|---|---|---|
| | | | | | (1661.5-15834.5) | (1176-14705.3) | | |
| sTNFR | - | - | - | - | 6356.4 (4039.8-10275.2) | 5558.4 (3542.8-9253.7) | 1.14 | 0.48 |
| MPO | - | - | - | - | 572165.9 (197021.2-1121886.2) | 345249.9 (186685.2-632967.7) | 1.66 | 0.09 |
| Ang-2 | - | - | - | - | 5478.9 (2858.6-10293.6) | 3912.7 (1779.8-7159.3) | 1.4 | **0.03** |

**Figure S1:** Plots of LCA model fit indicators in the LeoPARDS trial: a) Akaike Information Criterion (AIC); b) Bayesian Information Criterion (BIC) and c) log likelihood

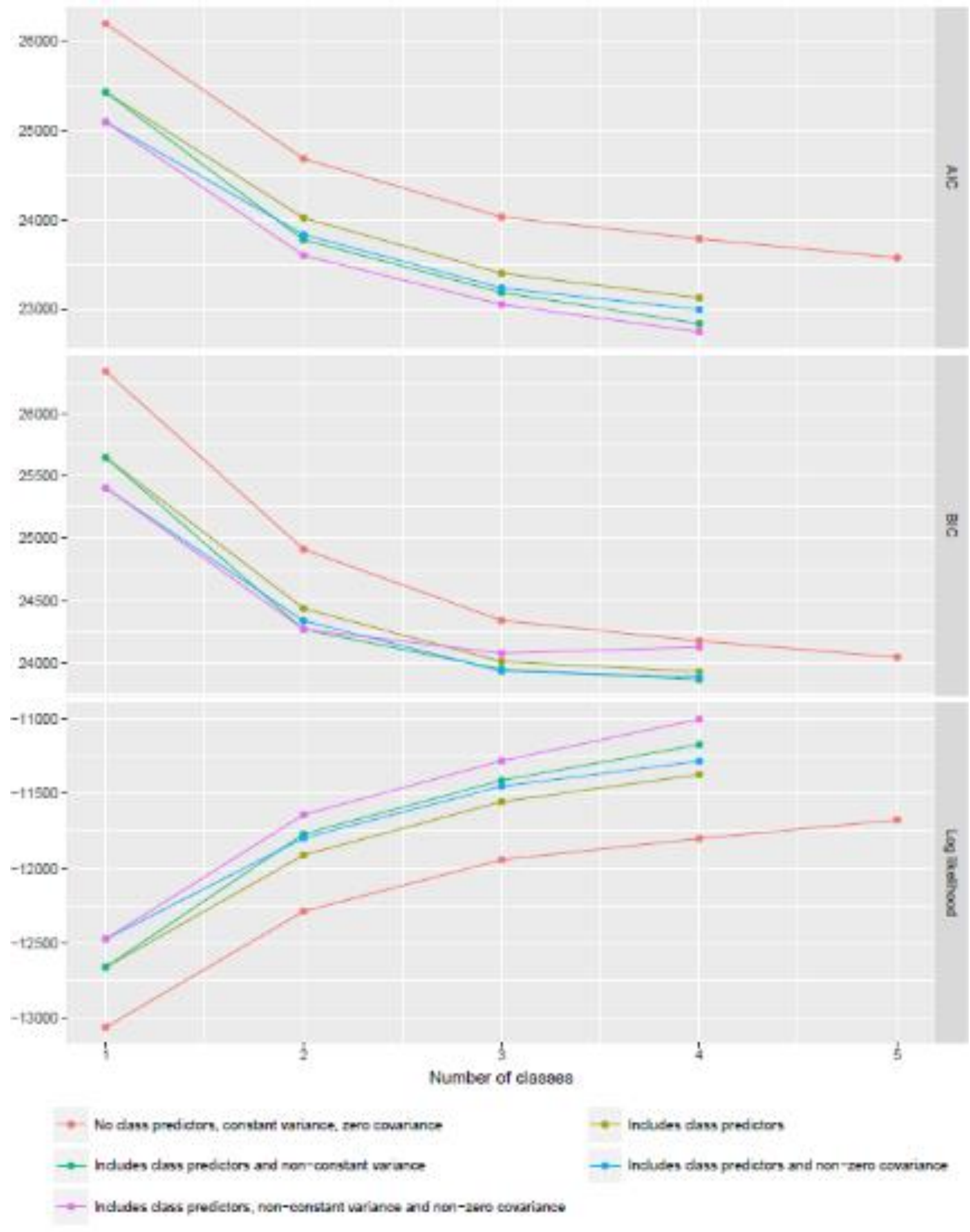

**Figure S2:** Class-specific sensitivity, specificity and c-statistics for multinomial logit models with increasing number of predictors, LeoPARDS trial

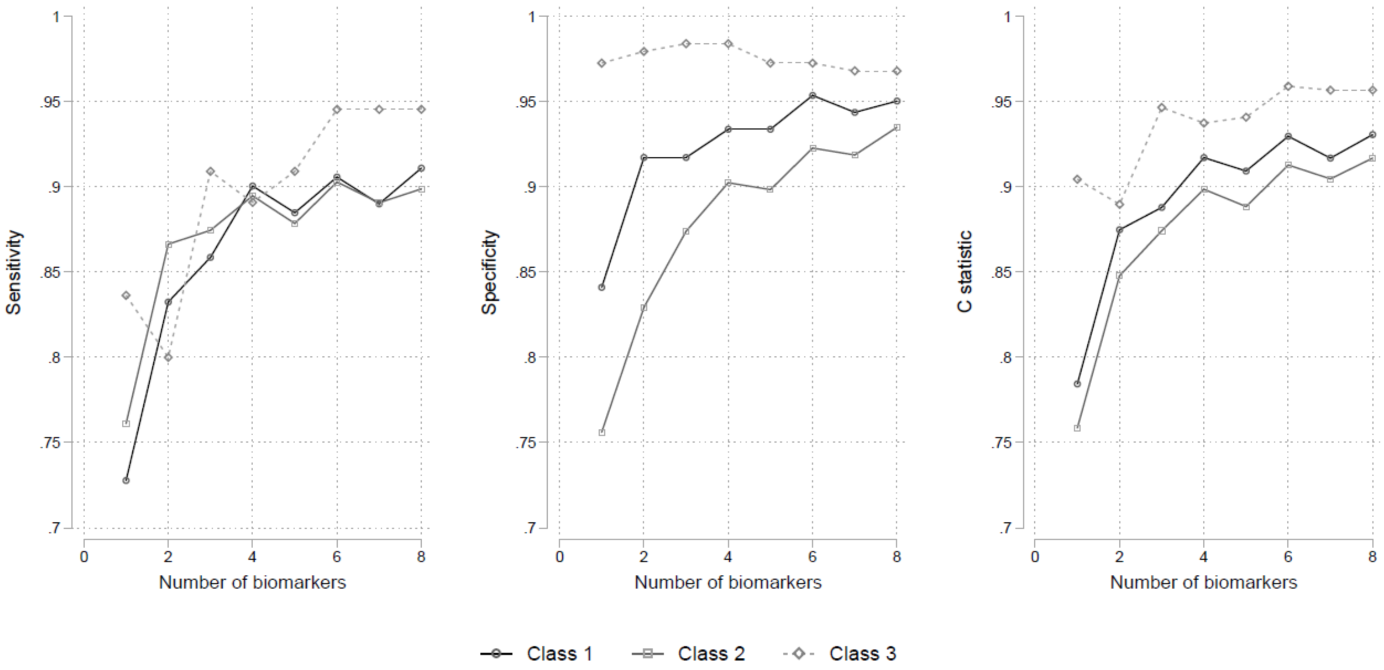

**Figure S2:** Plots of LCA model fit indicators in the VANISH trial: a) Akaike Information Criterion (AIC); b) Bayesian Information Criterion (BIC) and c) log likelihood

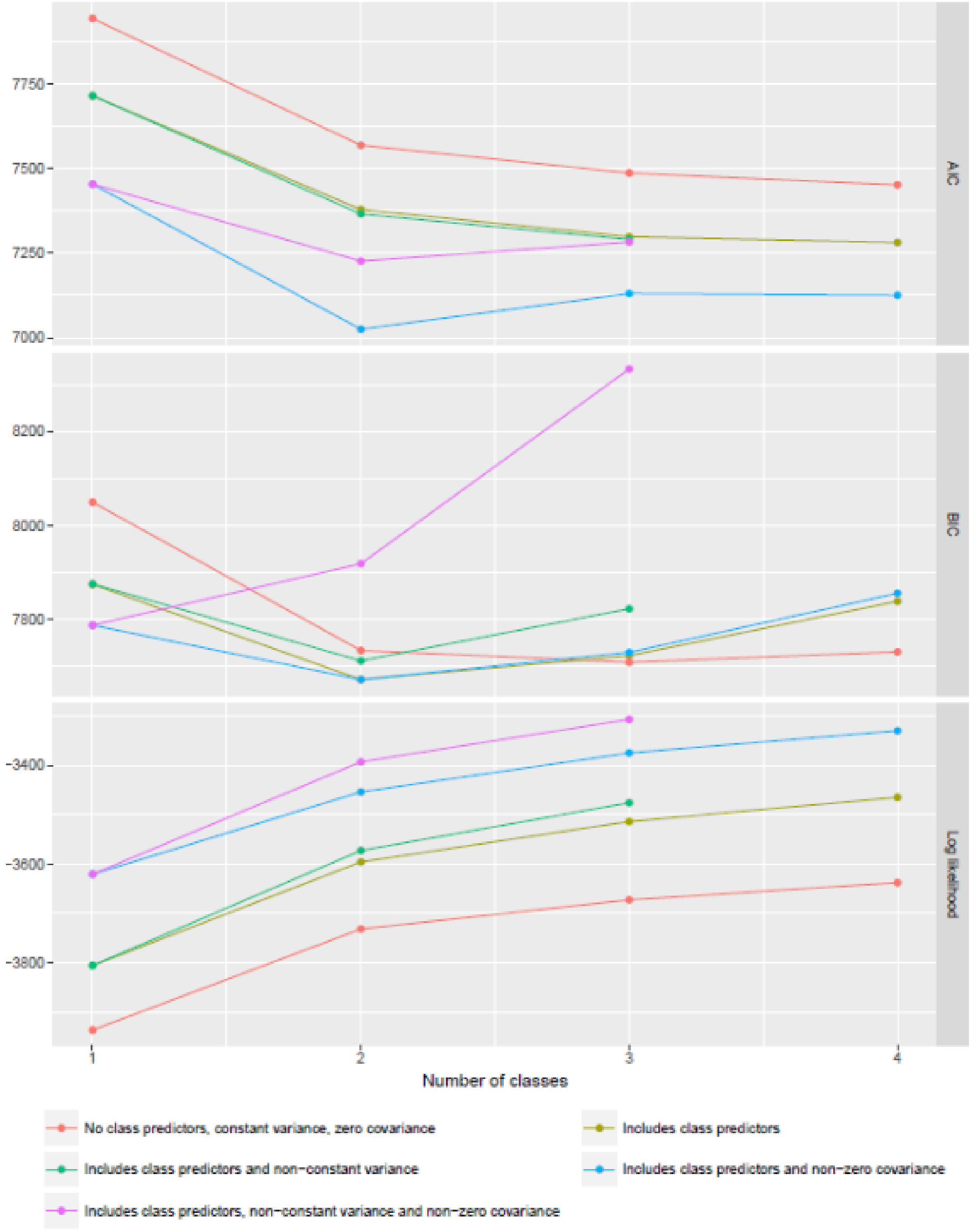

**Figure S4:** Assessment of hierarchical clustering performance in the full cytokine panels of GAinS (left), VANISH (middle) and LeoPARDS (right). (**A**) Test MSE (mean squared error) calculated by performing k-means clustering on 90% of samples and then taking the average of the squared distance of the 10% test samples to the closest cluster centers. 60 train/test sample sets were randomly drawn for each cluster number increasing from two to ten. (**B**) Heatmaps showing the level of consensus index which is the proportion of two samples which were in the same cluster during 1000 iterations following randomly resampling 80% of samples and 95% of mediators each time.

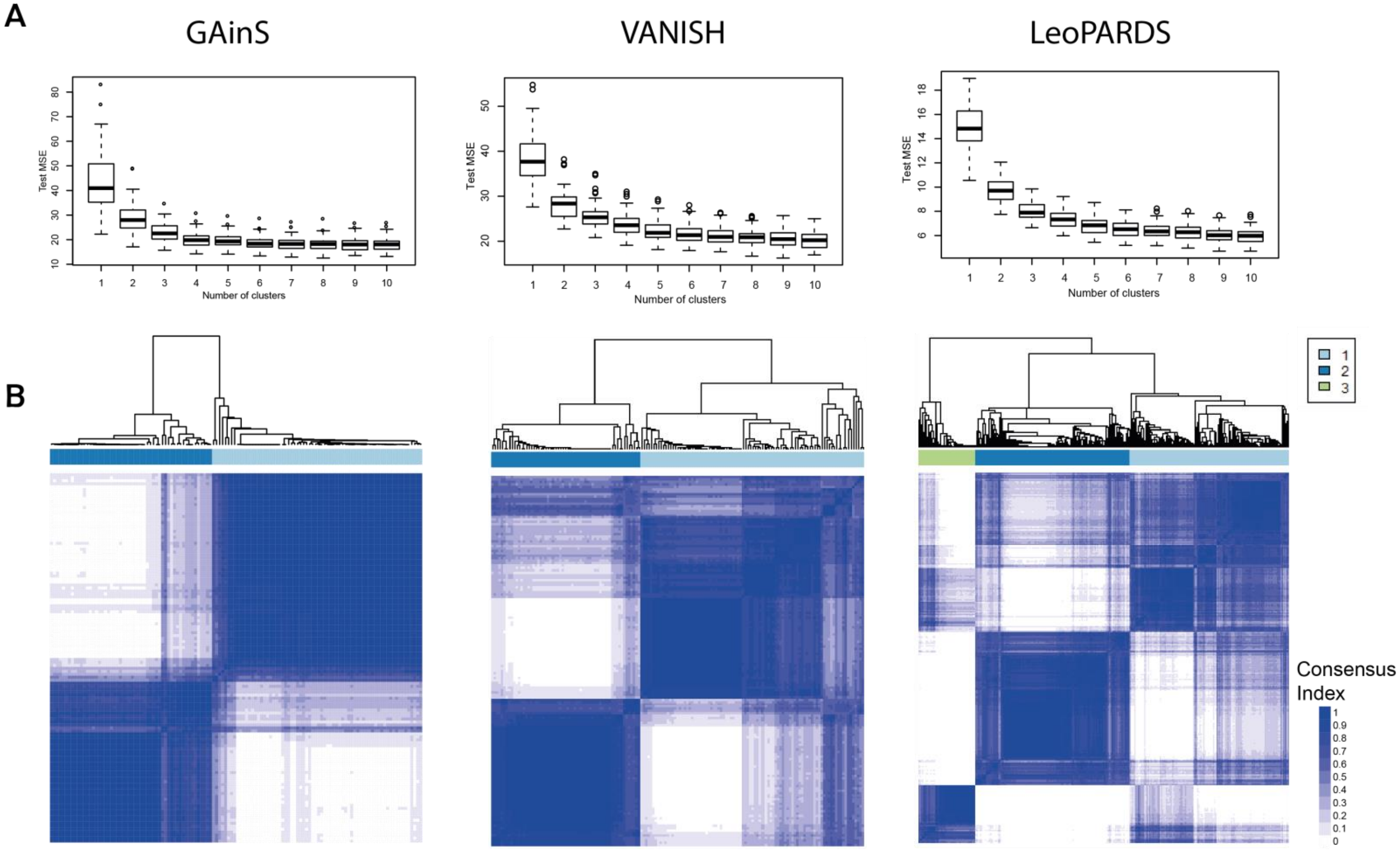

**Figure S5:** Hierarchical clustering of inflammatory mediators in baseline samples in GAinS. Heatmap colored by inflammatory mediator concentration, patients shown as columns (n=124) and mediators as rows (n=65), solid bars represent Sepsis Response Signature (SRS) assignment (light purple = SRS1, dark purple = SRS2, blank = no assignment available).

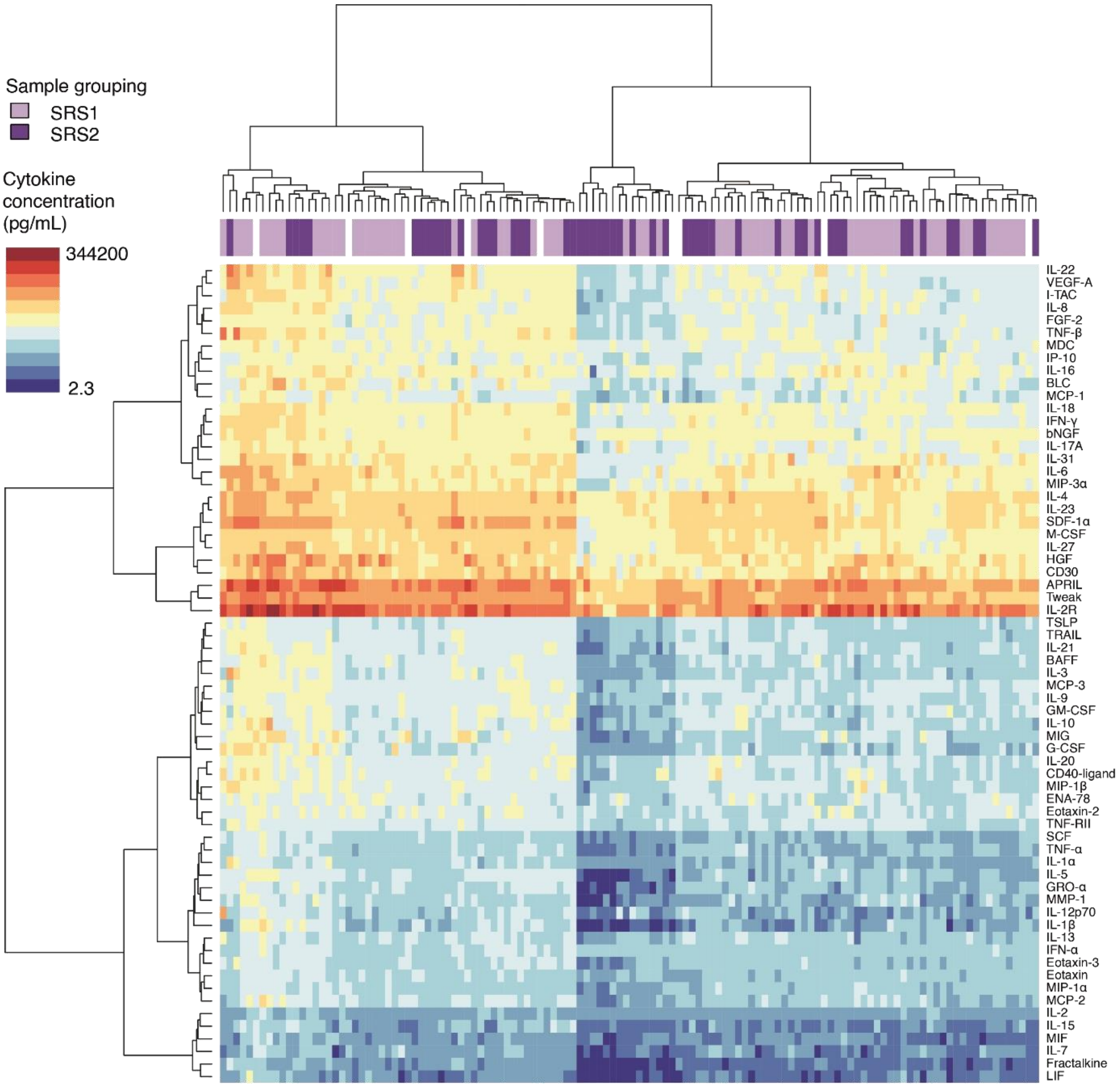

**Figure S6.** Comparison of 28-day mortality of patients in GAinS and VANISH stratified by SRS, cytokine clusters and a combination of the sub-phenotypes. In VANISH the analysis has been performed in all patients and in only those who were randomized to and received placebo as the second study drugs due to our previous findings of an interaction between SRS sub-phenotypes and use of hydrocortisone on 28-day mortality that could influence mortality when combining sub-phenotypes (23) (SRS1, sepsis response signature 1; SRS2, sepsis response signature 2; LC, low cytokine cluster, HC, high cytokine cluster; LC-SRS2 patients in both the low cytokine cluster and the SRS2 sub-phenotype). Red bars - GAinS, pale blue bars - VANISH all patients, dark blue bars – the placebo arm from VANISH.

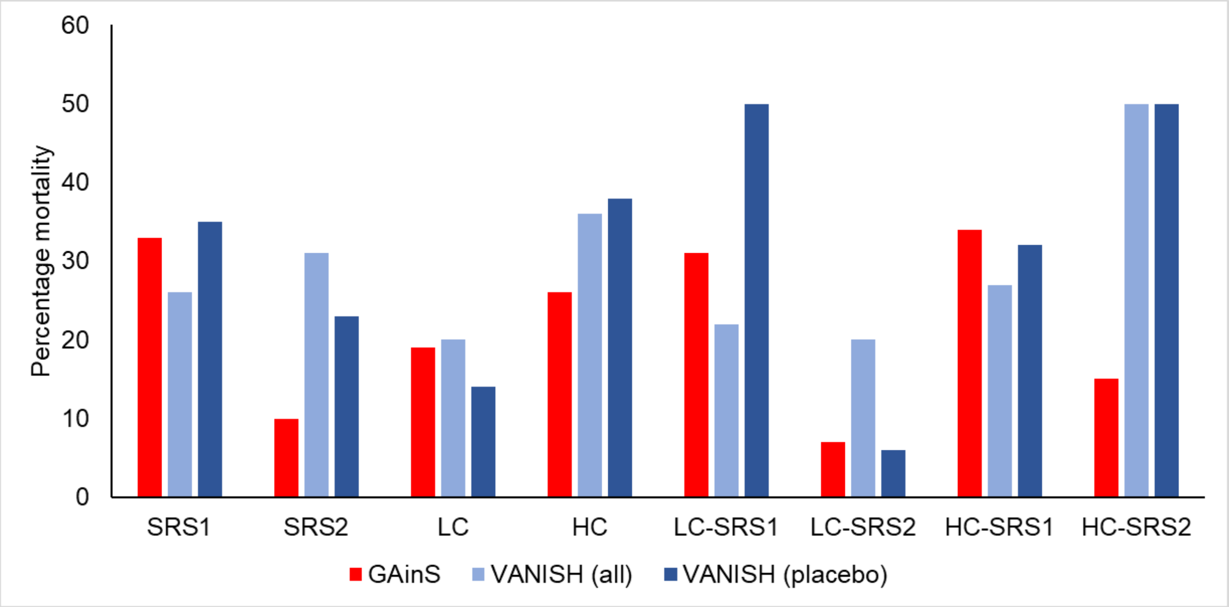

17. R Core Team. *R: a language and environment for statistical computing*. R Foundation for Statistical Computing; 2015.
